# Immersive Virtual Reality Training and Surgical Skill: A Systematic Review & Recommendations for Future Research

**DOI:** 10.1101/2024.06.17.24309027

**Authors:** Ami Gilliland, Erin Gaughan, Hannah Meek, Chandra Shekhar Biyani, Faheem Ijaz, George Gabriel, Ryan Mathew, Faisal Mushtaq

## Abstract

**Objectives:** In recent years, consumer-grade immersive virtual reality (iVR) systems have gained increasing attention for their potential applications in surgical training. The relatively low cost and increasing quality of these systems make them an appealing alternative to specialist surgical simulators, but their efficacy in comparison to traditional training techniques remains unclear. In this paper, we systematically review the recent literature comparing the impact of iVR-based and other training techniques on surgical proficiency.

**Method:** Five databases (Ovid MEDLINE, PsycINFO, EMBASE, Cochrane Library, Web of Science) were searched from 2016 to November 2023. 19 randomised controlled trials (totalling 593 participants) were identified as meeting the inclusion criteria for this review, involving trainee surgical participants being trained using iVR devices.

**Results:** Data from the 19 articles showed that iVR training was at least as effective as other simulation-based methods and more effective than conventional methods at improving key measures of surgical proficiency, including error rate, accuracy, and procedure-specific knowledge, with a short duration (20 minutes to 2 hours) being optimal. While these results demonstrate the potential of iVR-based training technologies to support effective and low-cost surgical skill training, the heterogeneity of the training tools and analysis methods used in the identified studies limits mechanistic explanations of the systems’ efficacy.

**Conclusions:** To support more robust and generalisable research into iVR surgical skill training, we make recommendations for the design and reporting of future intervention studies in this area. This notably involves the standardisation of an iVR definition, improvements to studies including consideration of personal experiences, and considering the long-term impacts of these interventions.

## 1 INTRODUCTION

Novice surgical trainees need to practice before they can operate on patients [1]. Traditionally, surgical training has adopted an opportunity-based apprenticeship model and has taken a “see one, do one, teach one” approach to learn surgical skills and procedures.[2],[3] This involves a surgical trainee observing and learning under the supervision of an experienced surgeon.[4] However, restrictions to working hours have meant that surgical trainees now have fewer opportunities to observe expert surgeons and practice surgical skills on live patients [5]. This issue was further exacerbated by the COVID-19 pandemic, which resulted in reduced patient access for surgical trainees.[6,7]

A potential solution to the challenges of training surgeons comes in the form of virtual reality (VR)-based simulation training. Simulation, in its broadest sense, allows surgical trainees to repeatedly practice surgical skills and procedures with no risk to patient safety.[8] Surgical trainees who receive simulation training have been found to display superior surgical skills when performing procedures on real patients.[9] VR surgical simulators are a relatively new method of simulation training. VR simulators use computer-generated graphics to mimic real-life surgical procedures, with some additionally having a visuo-haptic component which allows surgical trainees to practice the cognitive and sensorimotor skills specific to a procedure.[10–14]

Compared to basic simulators, such as benchtop anatomy models and box trainers, VR simulators have the advantage of being high-fidelity, a feature which has been associated with improved training outcomes.[11,12] They also allow immediate feedback, without requiring the presence of an expert surgeon.[12] Previous systematic reviews that have evaluated the effectiveness of VR as a surgical training tool conclude that VR simulators are as effective or more effective at improving the surgical skills of trainees compared to other training methods.[15–20] This suggests that VR simulators are a useful tool for surgical training and may reduce reliance on the traditional apprenticeship model. However, there are notable limitations. Specialist VR surgical simulators are expensive, with the costs of some exceeding $100,000, and lack versatility, with many designed to provide training on only one surgical procedure or group of procedures.[12,21]

Since 2016, there have been considerable advancements in VR technology with the introduction of commercially available immersive VR (iVR) devices which now offer a low-cost alternative to specialist VR simulators.[7,22,23] These devices are considered immersive as they use head-mounted displays (HMDs) to create a fully virtual interactive simulation.[24,25] iVR has the potential to increase the fidelity of surgical simulations as it is capable of creating an uninterrupted, fully scaled environment simulating many of the sensory stimuli present in a real-life operating room.[26] Whilst some specialist VR surgical simulators, such as the LapSim laparoscopic simulator, can be combined with HMDs to create similar immersive, high-fidelity simulations, commercially available iVR devices have the advantage of being more accessible due to their affordability, portability, and versatility.[25,27,28] These features of commercially available iVR devices mean that iVR is increasingly becoming a more viable training method for many surgical programmes.[7]

Numerous studies have examined the effectiveness of iVR as a surgical training method but have reported inconsistent results. For instance, Lohre et al.,[29] found that iVR training was more effective than traditional learning methods in teaching a complex surgical procedure. Conversely, Frederiksen et al. [28] concluded that iVR simulation results in poorer performance than conventional VR in laparoscopic training. A previous systematic review by Mao et al. [25] synthesised the results of these studies and found iVR training is more effective than other training methods. However, this review did not explain how characteristics of the iVR training, such as duration or surgical specialty, influence its effectiveness.

The primary objective of the present review is to investigate the effectiveness of immersive virtual reality (iVR) as a method of surgical training. To achieve this, the review seeks to answer several key questions from recently published randomised controlled trials (RCTs). Firstly, it aims to understand the impact of iVR training on various measures of surgical performance among surgical trainees. This involves assessing how trainees’ skills improve following iVR training and how these improvements compare to those achieved through other training methods. Secondly, the review explores how different characteristics of the iVR training intervention, such as its duration and the surgical speciality it targets, influence its effectiveness to allow for a more nuanced understanding of how iVR can be best utilized in different contexts. Thirdly, the review examines the effectiveness of iVR training across different levels of surgical trainees, from medical students to surgical residents, to determine whether the effectiveness of iVR training is modulated by skill and knowledge. Finally, the review provides a set of recommendations for design and reporting of future studies of iVR training, to help them support a more general explanation of the conditions under which iVR surgical training systems are effective.

## 2 METHODS

Randomised controlled trials (RCTs) published in the English language from 2016 onwards were eligible for inclusion. This date restriction was chosen as iVR is a relatively new technology, with a significant step-change in the availability of commercially available systems in recent years.[30] To be included in the study, RCTs had to meet specific inclusion criteria. The participants had to include individuals who were at a trainee surgical level, such as medical students, or residents. The intervention implemented in the study had to utilise an iVR device to provide training on either a surgical skill or procedure. An iVR device was defined as a device with a head-mounted display (HMD) that is used to present a computer-generated 3D, interactive interface. Mobile VR devices (e.g., Google Cardboard), high-end HMDs (e.g., Oculus Rift), and enhanced VR devices (e.g., HMDs combined with data gloves or bodysuits) were all considered to be iVR devices.[31] Additionally, specialist iVR devices specifically designed to provide surgical training were included if they had HMDs and a 3D interactive interface. Examples include the OssoVR and PrecisionOS.[32,33]

RCTs additionally had to include a comparator group that did not receive training using an iVR device, but no restrictions were placed on the type of comparator used. The main types of comparators used were conventional training methods, such as surgical videos, lectures, cadaver training, or traditional surgical guides. These comparator groups were used to address the objective concerning how iVR training compares to other types of surgical training methods in the synthesis.

Studies had to include an outcome measure related to participants’ subsequent performance on a task that required the surgical skills taught by the intervention to examine the transfer of learning from the virtual to the real-world environment. No restrictions were placed on what the outcome measure must be, but we grouped these measures into the following outcome domains in the synthesis to assess how iVR affects different measures of surgical proficiency. Our primary outcome domains included: (i) General Task Performance: Scores on general, quantitative measures of performance such as time to complete the procedure or error rate; (ii) Global Performance Ratings: Participants’ performance as rated by experienced surgeons on validated scales, such as the Objective Structured Assessment of Technical Skill (OSATS) [34]; (iii) Procedure-Specific Performance: Performance on assessment outcomes specifically related to the trained procedure or skill, such as the number of items completed on a procedure-specific checklist. Our secondary outcome measures included Experience Surveys assessing the subjective experience of using the immersive VR devices and other teaching methods included as a control.

### 2.1 Selection Process & Data Extraction

Five databases (Ovid MEDLINE, PsycINFO, EMBASE, Cochrane Library, Web of Science) were searched from 2016 to 16/11/23. Search strategies were based on the PICO framework. The terms “randomly, randomized, intervention, and double-blind” were included in the search strategy as these terms have been found to have the greatest specificity when searching databases for RCTs.[35] “Randomized” was modified to “randomised” to increase sensitivity due to variations in American and English spelling. All searches had limits applied to exclude reports that were published prior to 2016 or were not published in the English language, as per the eligibility criteria. Additionally, as only published RCTs were eligible for inclusion, limits were applied to exclude other types of records. The final search strategy was peer-reviewed using the Peer Review of Electronic Search Strategies (PRESS) checklist [36] by an independent researcher, who deemed it to be clear and comprehensive.

For the first run of searches (completed from 2016 to 11/11/21), five researchers were separated into two sets of pairs, with one individual (author EG) serving to review inconsistencies. The study selection was completed using double screening to improve reliability. The following Systematic Review Accelerator Tools were used: “Deduplicator”, “Screenatron”, “Disputatron”.[37]. Records identified through database and citation searching were imported into “Deduplicator” and records identified as duplicates were checked by author AG before being excluded. Following this, the overall number of retrieved articles (825) was divided equally, and each pair was allocated one subsection (412 papers for pair one, 413 papers for pair two). The two researchers independently screened titles using a standardized title and abstract form, “Screenatron”. In cases of disagreement, consensus on which articles to include was decided by the reviewer, EG, using a tool that detects screening disagreements and decisions between reviewers (“Disputatron”). Next, the same sets of pairs independently screened the full-text articles for inclusion. Full texts were retrieved using the Endnote “Find Full Text” feature. Full texts which Endnote failed to find were manually retrieved by the researchers and any full texts which could not be retrieved were excluded from the review. Similarly, in cases of disagreement, a consensus was reached on inclusion or exclusion by the reviewer EG using Disputatron. Due to the lack of clarity in many of the reports about the type of VR used in the study, reports that passed this stage of eligibility screening were additionally screened by an expert in iVR (FM) to ensure they met the criteria for iVR.

For the re-run of the searches (completed from 2021 to 16/11/23), one researcher used Systematic Review Accelerator Tools (“Deduplicator” and “Screenatron”) to screen the studies. As before, records identified through searching the databases were imported into “Deduplicator”, records identified as duplicates were checked by author HM before exclusion. The total number of retrieved articles (419) were then screened using “Screenatron” through reviewing titles and abstracts. Subsequently, the full-text articles were screened for inclusion. The texts were retrieved manually, and any that could not be retrieved were excluded from the review.

For the initial run of searches, data was extracted using the standard data collection form for intervention reviews, created by the Cochrane Collaboration.[38] For each eligible study, two review authors independently completed a data extraction form. Extracted data were then compared by a third reviewer, who resolved any discrepancies between data collectors by sourcing the accurate information from the original publication to create a final data extraction form for each included report.

Means and standard deviations for each group, as well as p-values, were also recorded for each outcome measure. For studies that did not report standard deviations, where possible, standard deviations were calculated from the confidence intervals using the Cochrane guidelines.[39] Following the Synthesis Without Meta-analysis (SWiM) reporting guidelines,[40] the synthesis method chosen was vote counting based on direction of effect. This was conducted by categorizing each effect estimate as showing either benefit or harm based on the observed direction of effect, thereby creating a standardised binary metric to aid comparisons.[41] This approach was selected because it became clear that there was no consistent effect measure reported across studies.

## 3 RESULTS

### 3.1 Study Selection & Characteristics

In total, 1393 records were identified through database searching from 2016 to 16/11/2023, and an additional 63 were identified through citation searching of a relevant review. [42] Following the removal of duplicates (149 records), 1244 titles and abstracts were screened at the record screening stage. For the initial run of searches (2016- to 11/11/2021) which screened 825 titles and abstracts, the screening agreement between pairs at this stage was 78% for pair 1 (AG and GG) and 71% for pair 2 (LH and MC). Following record screening, the full texts of 190 records were retrieved and screened to determine whether they met the full eligibility criteria. At this stage of screening, the screening agreement was 79% for pair 1 (AG & GG) and 57% for pair 2 (LH and MC). For the re-run of searches (2021-16/11/2023), 419 titles and abstracts were screened, of which the full texts of 17 records were retrieved and screened to determine if they met the eligibility criteria. Of the 207 full texts that were screened, 169 were excluded for the following reasons: they were duplicates which had not been identified by the Deduplicator tool (n=47), the participants were not surgical trainees (n=4), the intervention did not meet the criteria for iVR (n=75), the study was a review or did not meet the criteria to be considered an RCT (n=36), none of the primary outcome measures were assessed (n=8), the study did not have the necessary control group (n=1). Overall, 19 studies met our eligibility criteria. The key characteristics of these studies are presented in Table 1. Table 2 provides additional details about the iVR simulator used in each included study.

**Table 1:** Study Characteristics.

| Study | Participants | Procedure/Skill | Outcome Assessment | Outcome Measures |
| --- | --- | --- | --- | --- |

**Table 1: Study Characteristics**
|  |  |  |  | **General Task**<br>**Performance** | **Global**<br>**Performance** | **Procedure-Specific** | **Surveys** |
| --- | --- | --- | --- | --- | --- | --- | --- |
| Abulfaraj et al. (2021) | 42 (intervention = 22, control = 20) medical interns | Managing anaphylaxis and status epilepticus | Managing status epilepticus on a low-fidelity mannequin | Time-to-critical actions | - | - | Subjective confidence |
| Andersen et al. (2021) | 19 (intervention = 10, control = 9) medical student | Ultrasound-guided PVC placement | Placing 3 PVCs in a phantom using ultrasound guidance | Time to complete taught skills | - | Proportion of successful cannulations and surface punctures | - |
| Blumstein et al. (2020) | 20 (intervention = 10, control = 10) medical students | Tibial shaft fracture intramedullary nailing | Simulated tibial shaft fracture intramedullary nailing procedure | Time to complete procedure | Global Assessment 5-point Rating Scale | Procedure-Specific Checklist | - |
| Cevellos et al. (2022) | 20 (intervention = 10, control = 10) medical students and residents | Pinning of a slipped capital femoral epiphysis (SCFE) | Placing a guidewire in a SawBones SCFE model (realistic artificial proximal femur of a moderate SCFE) using fluoroscopic guidance | Total time-to-pin placement | - | Number of "in-and-outs"<br>Number of fluoroscopy images taken<br>Number of articular surface penetrations<br>Angle between the pin and the physis<br>Pin-tip location with respect to centre-centre position<br>Pin tip location with respect to distance from subchondral bone | - |
| Crockatt et al. (2023) | 14 (intervention = 6, control = 8) junior residents | Reverse total shoulder arthroplasty (rTSA) | Demonstrate key steps involved in implantation of the Comprehensive Augmented Baseplate | Time to completion of the baseplate implantation | Objective Structured Assessment of Technical Skill (OSATS) checklist<br>Global Rating Scale (GRS) | Pre and post training written knowledge scores (%) | - |
| Frederiksen et al. (2020) | 31 (intervention = 16, control = 15) first-year residents | Laparoscopic salpingectomy | Performance on either the conventional VR or immersive VR simulators (depending on group) | Cognitive load (secondary task RT)<br>Time to complete procedure<br>Blood loss<br>Path length<br>Angular path<br>Number of major vessels cut | - | Diathermy damage<br>Distance tube cut from the uterus | Motion sickness |
| Gomindes et al. (2023) | 31 surgical trainees (numbers in groups not stated) | Insertion of a short cephalomedullary TFNA | Insert short cephalomedullary TFNA systema into dry saw-bone model femurs | Time to complete procedure | - | Tip-apex distance (TAD)<br>Number of lag-screw and entry guidewire insertion attempts | - |
| Lamb et al. (2023) | 38 (intervention = 19, control = 19) medical students | Tibia intramedullary nail (IMN) surgery | Simulation of intramedullary nail insertion on model tibia bones | Time to complete procedure | Global Assessment 5-point Rating Scale | Procedure-Specific Checklist | Pre and post surveys measuring experience |
| Lohre et al. (2020a) | 18 (intervention = 9, control = 9) senior orthopedic surgery residents | Reverse shoulder arthroplasty | Augmented glenoid implantation on a cadaver | Time to complete procedure<br>Error rate | Objective Structured Assessment of Technical Skills (OSATS) | Knowledge Test Scores | - |
| Lohre et al. (2020b) | 19 (intervention = 9, control = 10) orthopedic surgical residents | Glenoid exposure | Glenoid exposure on a cadaver | Time to complete procedure | OSATS | Knowledge Test Scores | - |
| Margalit et al. (2022) | 21 (VR intervention = 7, PS intervention = 7, control = 7) orthopaedic trainees | Slipped capital femoral epiphysis model | In-situ pin fixation on the phantom limb with conventional C-Arm | Overall surgical procedure time | Global Rating Scale (GRS) score | Number of radiographs taken<br>Radiographic screw position<br>Physical screw accuracy<br>Presence of breeching of articular surface of femoral neck<br>Life like rating of physical module<br>Overall platform rating | - |
| McKinney et al. (2022) | 22 (intervention = 11, control = 11) orthopaedic surgery residents | Fixed bearing medial unicompartmental knee arthroplasty (UKA) | Medial UKA on SawBones model | Timed for training length and procedure completion | Global Assessment 5-point Rating Scale | Procedure specific checklist | Pre and post surveys measuring experience |
| Mok et al. (2021) | 121 (intervention = 60, control = 61) medical students | Tendon repair | Tendon repair on a synthetic model | - | Global Rating Scale (GRS) | - | - |
| Ros et al. (2021) | 89 (intervention = 45, control = 44) medical students | Lumbar puncture | Oral knowledge assessment and lumbar puncture on mannequin | Time to complete procedure | - | Knowledge Test Scores<br>Procedure-Specific Checklist | - |
| Tapiala et al. (2023) | 30 (intervention = 16, control = 14) medical students | Mastoidectomy | Simulated mastoidectomy on 3D printed temporal bone models | Total operational time | - | Timestamps of completed subtasks and requests for assistance<br>Evaluation of drilled temporal bones by experts using modified Welling Scale | Self-assess performance |
| Wan et al.<br>(2023) | 20 (intervention = 10, control = 10) medical students | Orthognathic surgery | Simulated bimaxillary orthognathic surgery | Operation time<br>Timeouts | - | Number of instrument selection errors<br>Number of position and angular errors<br>Number of prompts required to progress to next step | - |
| Xin et al.<br>(2019) | 16 (intervention = 8, control =8) surgical graduate students | Pedicle screw placement | Pedicle screw placement on cadaver | Time to complete procedure<br>Accuracy<br>Success Rate | - | - | - |
| Yogonathan et al. (2018) | 29 (intervention = 17, control = 12) foundation year doctors | Single handed reef knot | Ability to tie a single-handed reef knot | Time to complete procedure | Rating on marking criteria developed by Royal College of Surgeons | - | - |
| Zaid et al.<br>(2022) | 22 (intervention = 11, control = 11) medical students or residents | Unicompartmental knee arthroplasty | Unicompartmental knee arthroplasty on SawBones | Time to complete procedure | OSATS | - | - |

**Table 2:**
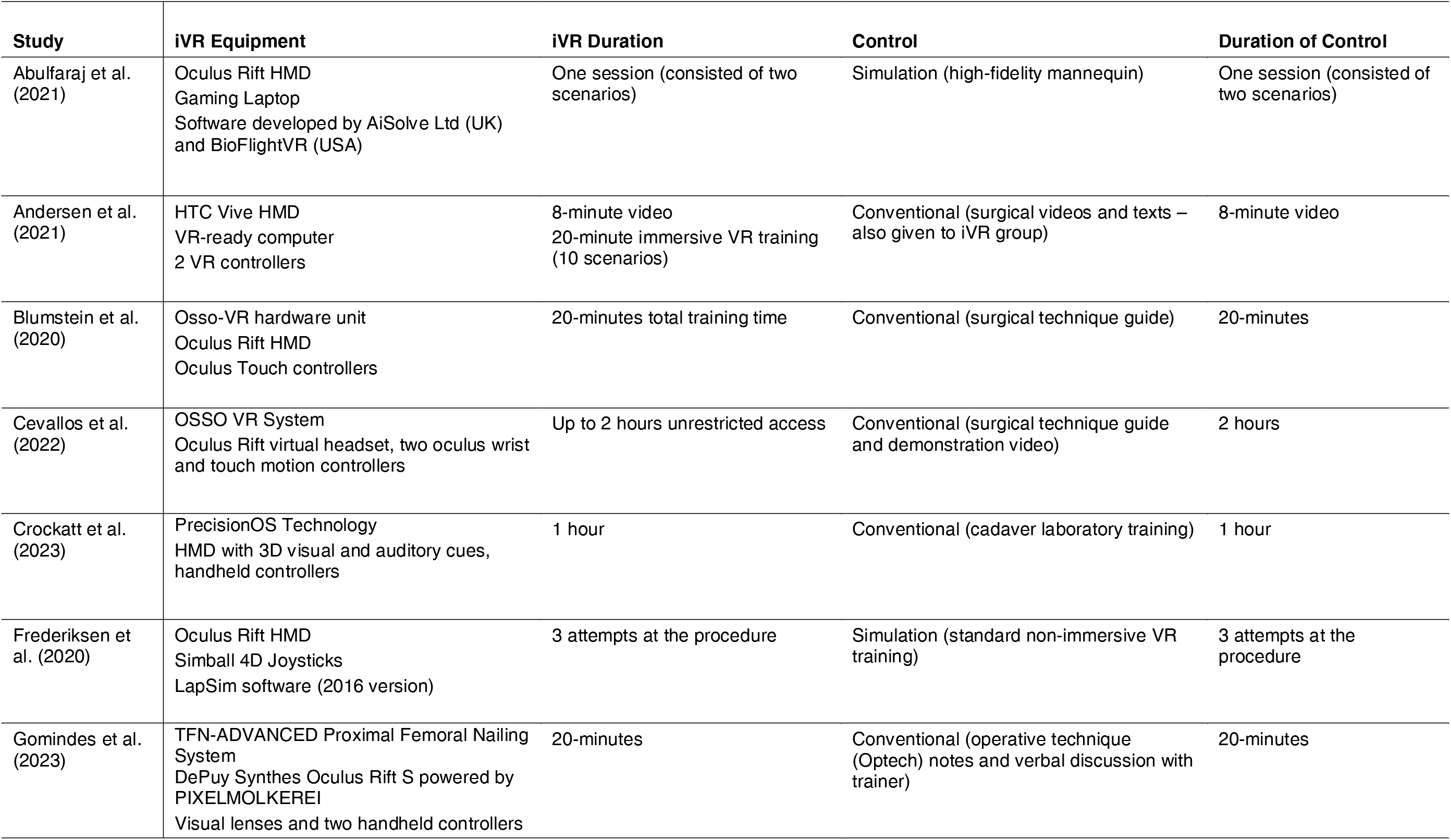
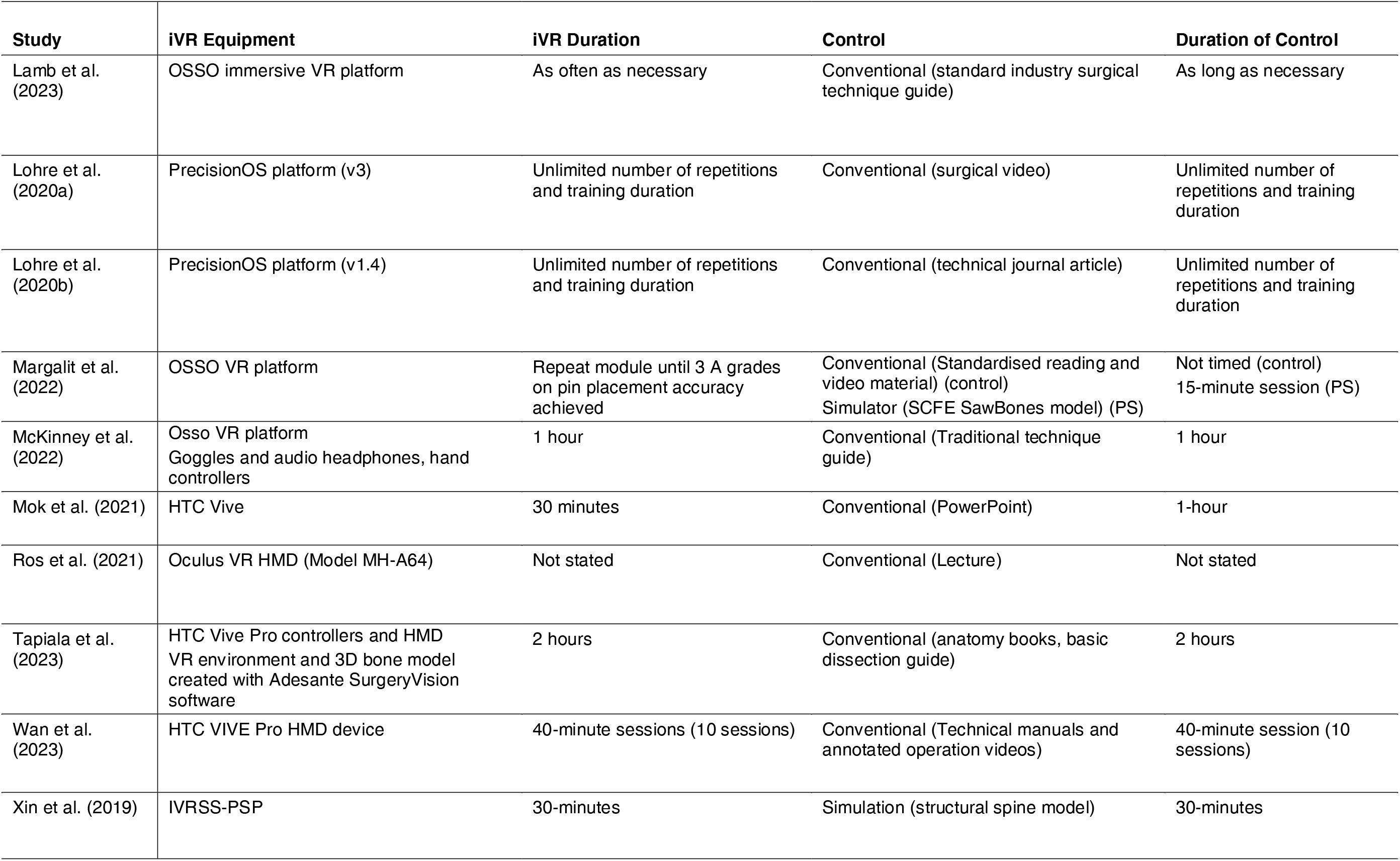
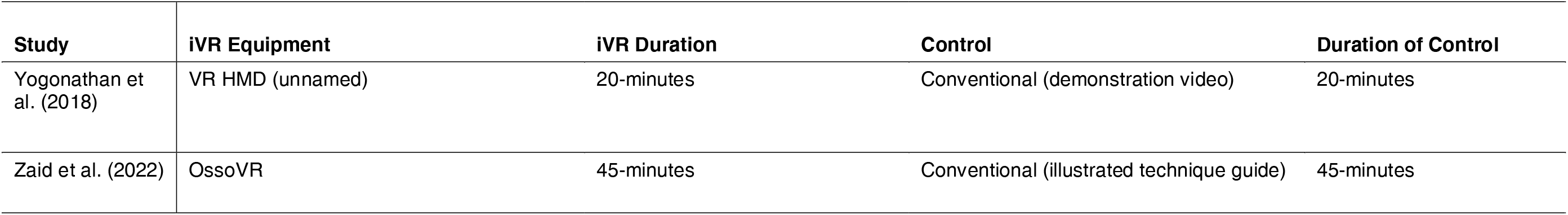
iVR and comparator interventions used in each included study.

| **Study** | **iVR Equipment** | **iVR Duration** | **Control** | **Duration of Control** |
| --- | --- | --- | --- | --- |
| Abulfaraj et al. (2021) | Oculus Rift HMD<br>Gaming Laptop<br>Software developed by AiSolve Ltd (UK) and BioFlightVR (USA) | One session (consisted of two scenarios) | Simulation (high-fidelity mannequin) | One session (consisted of two scenarios) |
| Andersen et al. (2021) | HTC Vive HMD<br>VR-ready computer<br>2 VR controllers | 8-minute video<br>20-minute immersive VR training (10 scenarios) | Conventional (surgical videos and texts – also given to iVR group) | 8-minute video |
| Blumstein et al. (2020) | Osso-VR hardware unit<br>Oculus Rift HMD<br>Oculus Touch controllers | 20-minutes total training time | Conventional (surgical technique guide) | 20-minutes |
| Cevallos et al. (2022) | OSSO VR System<br>Oculus Rift virtual headset, two oculus wrist and touch motion controllers | Up to 2 hours unrestricted access | Conventional (surgical technique guide and demonstration video) | 2 hours |
| Crockatt et al. (2023) | PrecisionOS Technology<br>HMD with 3D visual and auditory cues, handheld controllers | 1 hour | Conventional (cadaver laboratory training) | 1 hour |
| Frederiksen et al. (2020) | Oculus Rift HMD<br>Simball 4D Joysticks<br>LapSim software (2016 version) | 3 attempts at the procedure | Simulation (standard non-immersive VR training) | 3 attempts at the procedure |
| Gomindes et al. (2023) | TFN-ADVANCED Proximal Femoral Nailing System<br>DePuy Synthes Oculus Rift S powered by PIXELMOLKEREI<br>Visual lenses and two handheld controllers | 20-minutes | Conventional (operative technique (Optech) notes and verbal discussion with trainer) | 20-minutes |
| Lamb et al. (2023) | OSSO immersive VR platform | As often as necessary | Conventional (standard industry surgical technique guide) | As long as necessary |
| Lohre et al. (2020a) | PrecisionOS platform (v3) | Unlimited number of repetitions and training duration | Conventional (surgical video) | Unlimited number of repetitions and training duration |
| Lohre et al. (2020b) | PrecisionOS platform (v1.4) | Unlimited number of repetitions and training duration | Conventional (technical journal article) | Unlimited number of repetitions and training duration |
| Margalit et al. (2022) | OSSO VR platform | Repeat module until 3 A grades on pin placement accuracy achieved | Conventional (Standardised reading and video material) (control)<br>Simulator (SCFE SawBones model) (PS) | Not timed (control)<br>15-minute session (PS) |
| McKinney et al. (2022) | Osso VR platform<br>Goggles and audio headphones, hand controllers | 1 hour | Conventional (Traditional technique guide) | 1 hour |
| Mok et al. (2021) | HTC Vive | 30 minutes | Conventional (PowerPoint) | 1-hour |
| Ros et al. (2021) | Oculus VR HMD (Model MH-A64) | Not stated | Conventional (Lecture) | Not stated |
| Tapiala et al. (2023) | HTC Vive Pro controllers and HMD<br>VR environment and 3D bone model created with Adesante SurgeryVision software | 2 hours | Conventional (anatomy books, basic dissection guide) | 2 hours |
| Wan et al. (2023) | HTC VIVE Pro HMD device | 40-minute sessions (10 sessions) | Conventional (Technical manuals and annotated operation videos) | 40-minute session (10 sessions) |
| Xin et al. (2019) | IVRSS-PSP | 30-minutes | Simulation (structural spine model) | 30-minutes |

**Table 2:** iVR and comparator interventions used in each included study.
| Study | iVR Equipment | iVR Duration | Control | Duration of Control |
| --- | --- | --- | --- | --- |
| Yogonathan et al. (2018) | VR HMD (unnamed) | 20-minutes | Conventional (demonstration video) | 20-minutes |
| Zaid et al. (2022) | OssoVR | 45-minutes | Conventional (illustrated technique guide) | 45-minutes |

### 3.2 Results of Individual Studies

Twelve of the nineteen included studies concluded that participants who received iVR training performed better on measures of surgical performance than participants in the comparator group. Six studies concluded that there was no difference in surgical performance following training via iVR and another method. Only one study [28] concluded that iVR training led to worse surgical performance. For each outcome measure, summary statistics from each of the included studies are displayed in Table 3, including the mean (M), standard deviation (SD), and sample size (N) of each group, as well as the resulting p-value. Where it was not possible to calculate or retrieve any values, it has been left blank.

**Table 3:**
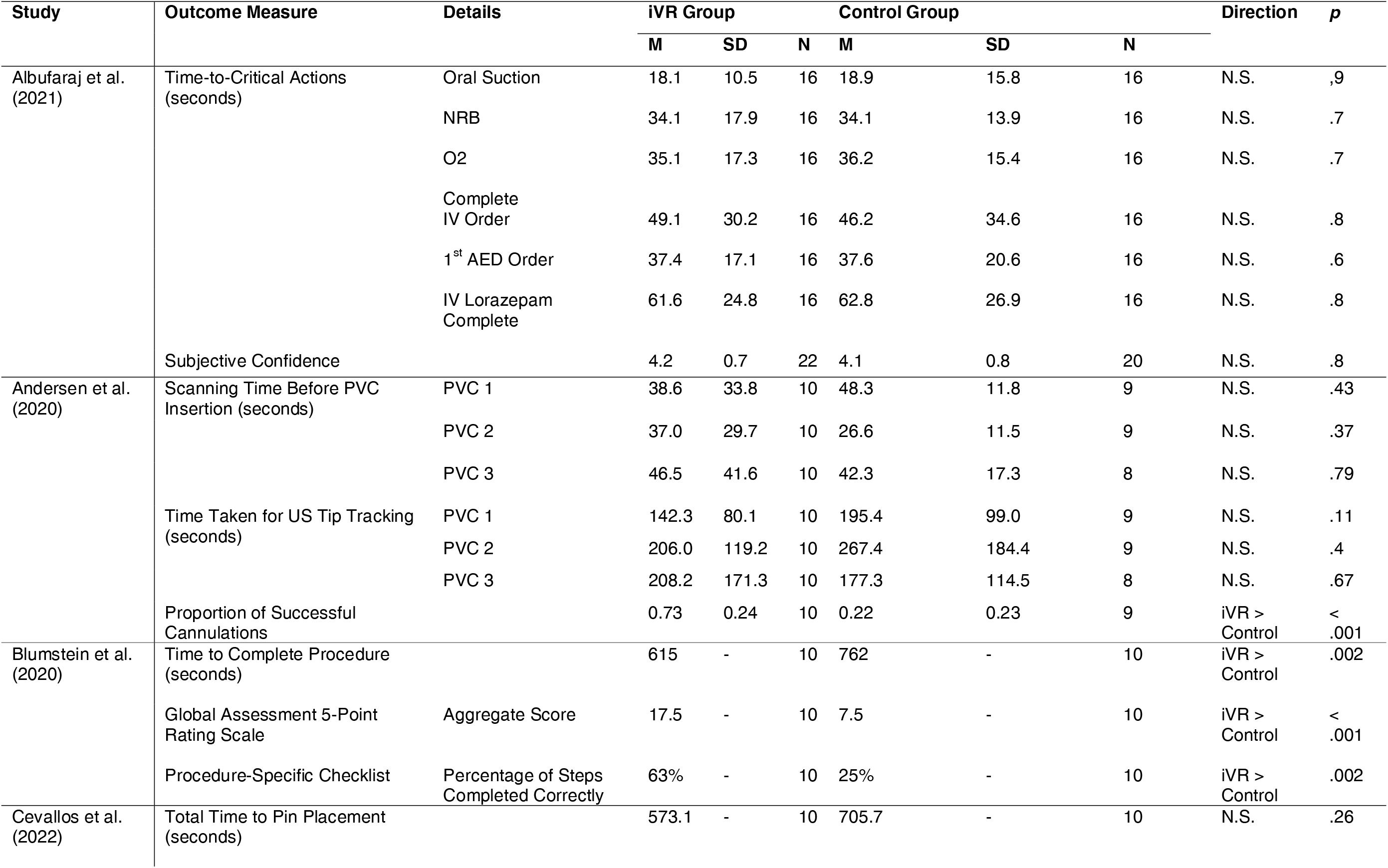
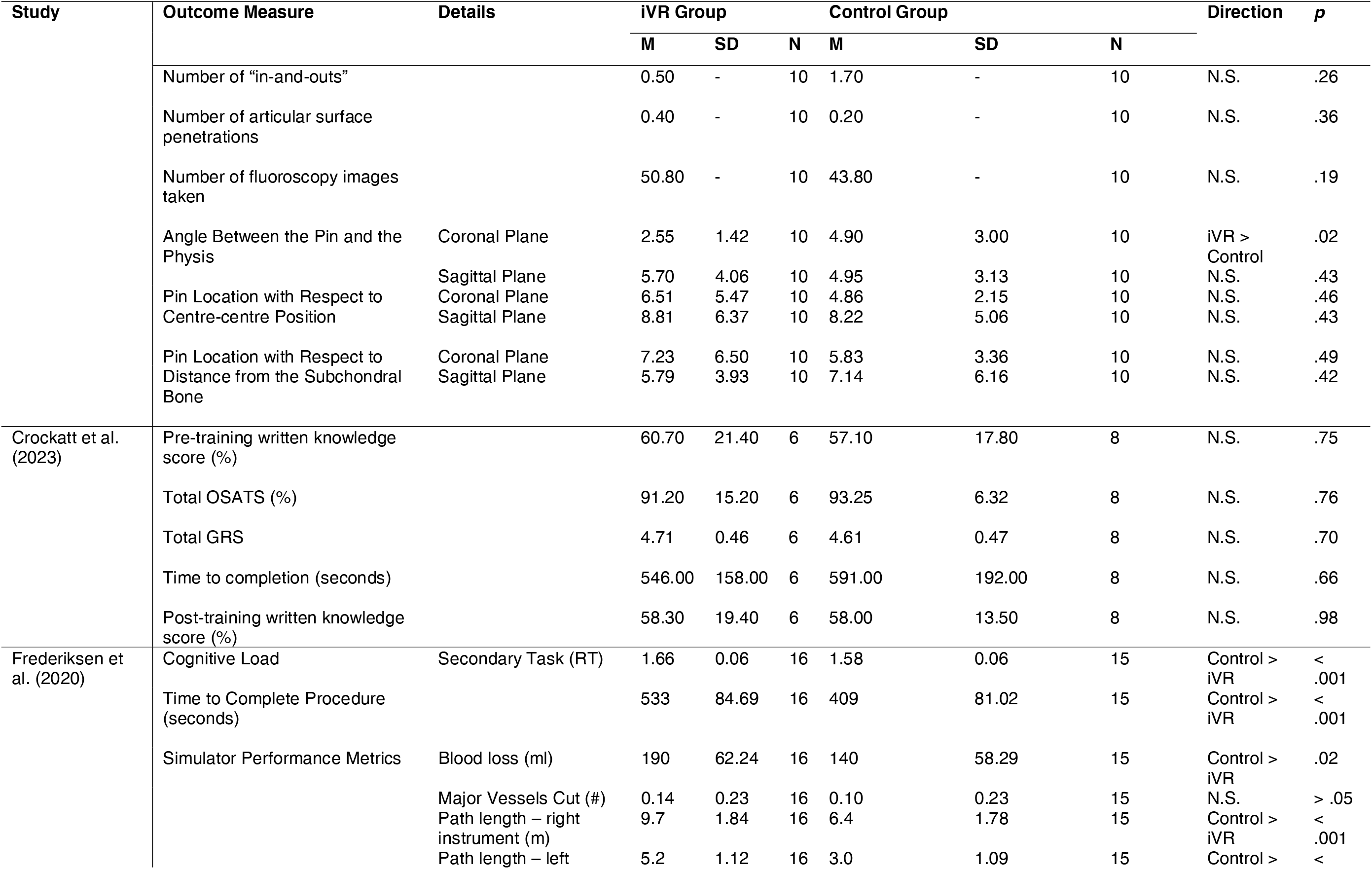
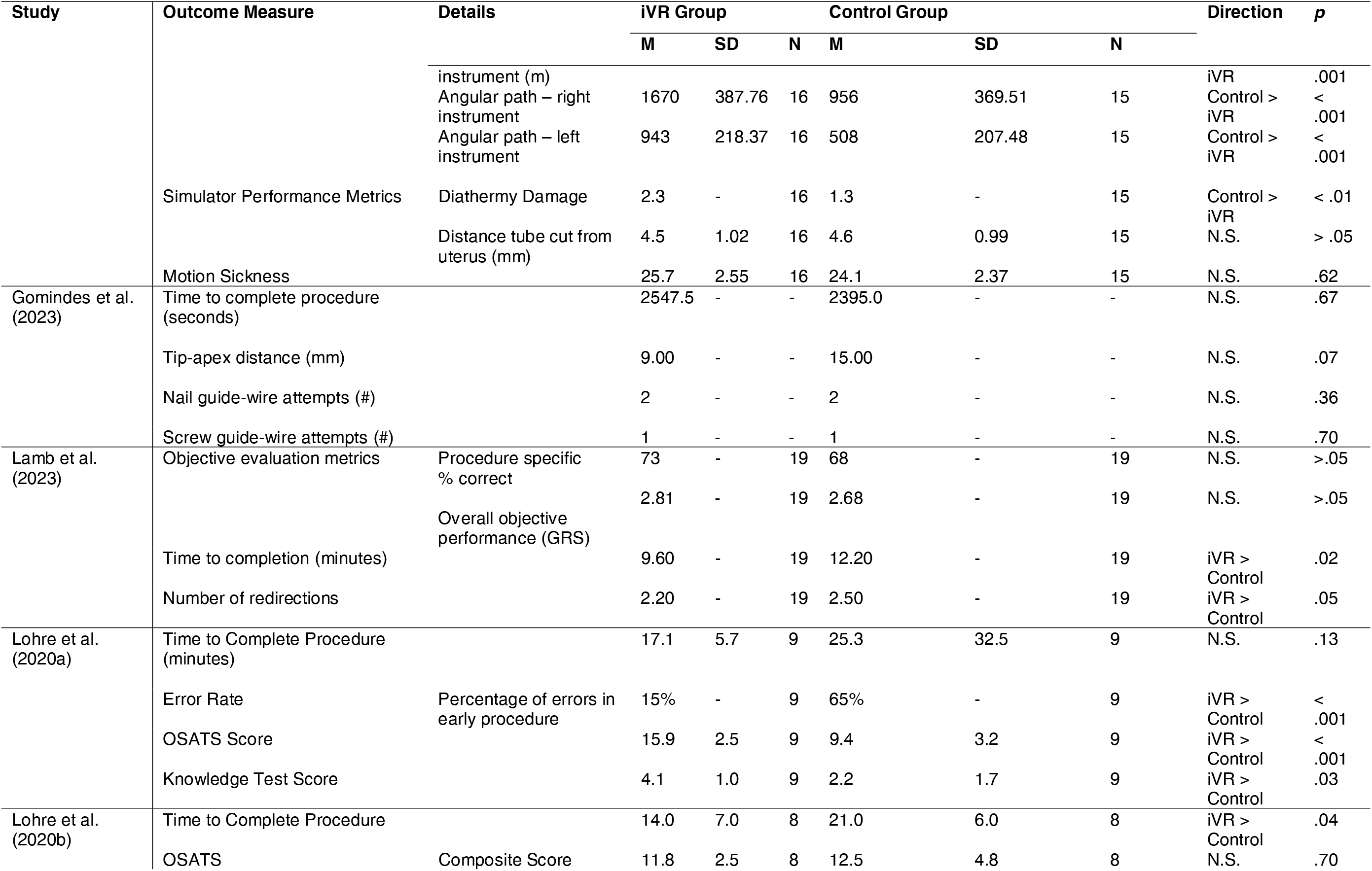
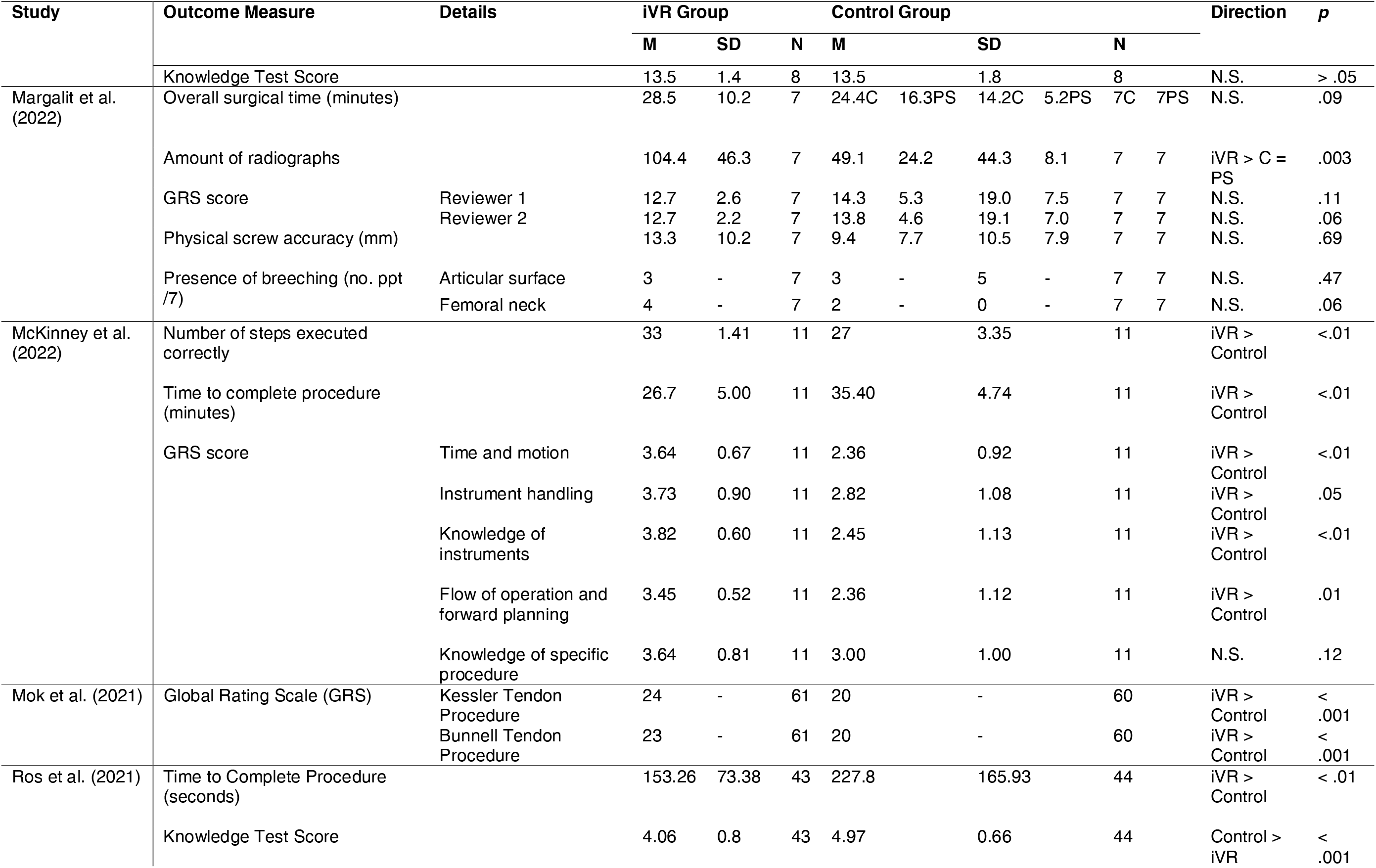
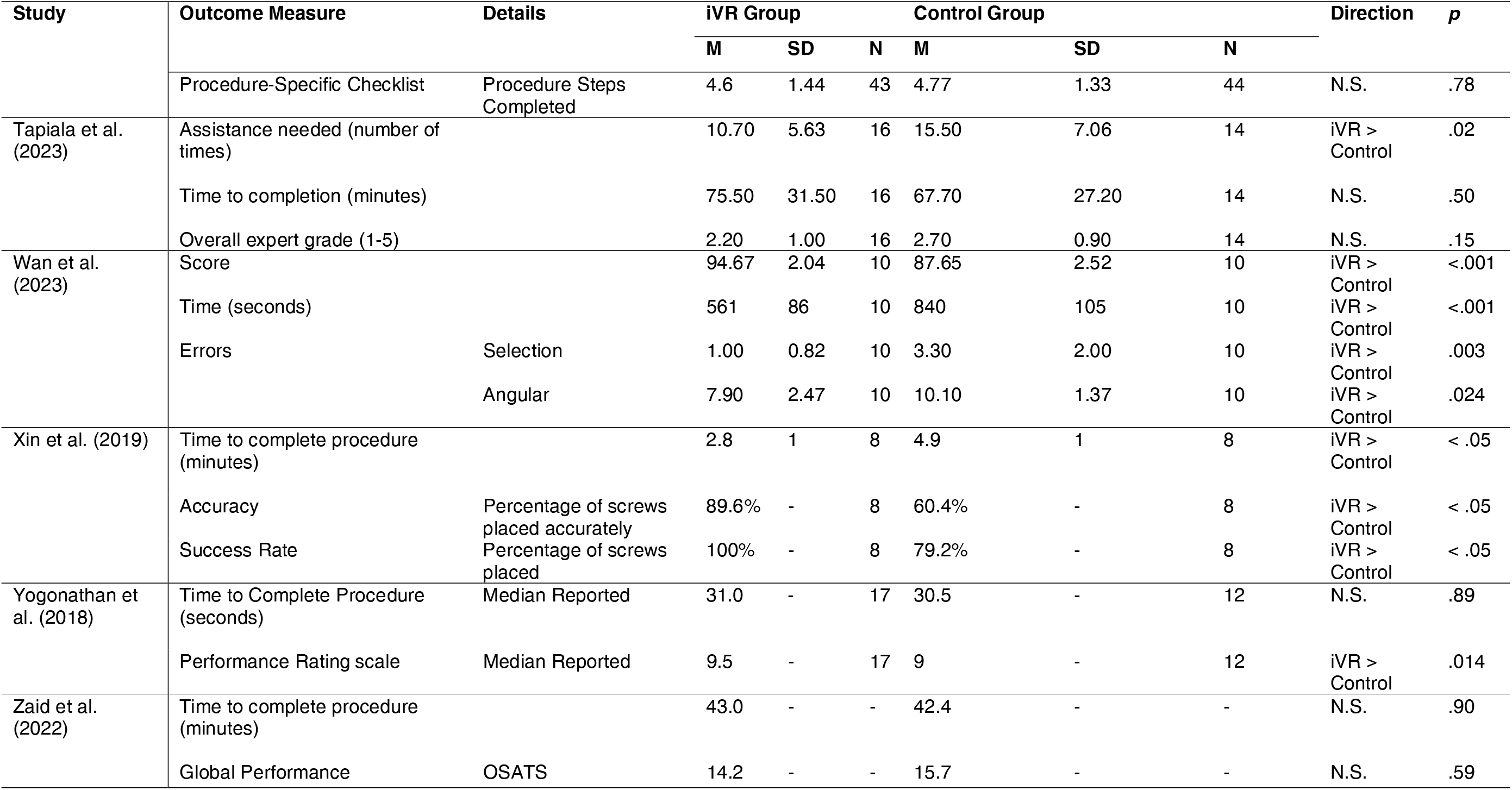
Results of individual included studies.

### 3.3 Results of Synthesis

To explore the effects of iVR training on different measures of surgical proficiency, studies were grouped based on the outcome domains outlined in the methods section (see Table 4). All outcome measures related to participants’ performance following training, when they were assessed whilst performing the taught procedure or skill. Experience surveys were not included in the synthesis, as only four studies had reported this outcome and the surveys used concerned different topics (confidence, motion sickness, general VR experience, and surgical VR experience). To evaluate whether the reported effects of iVR training differed between studies that used different comparator groups, studies were additionally grouped according to the comparator (Table 4). Fifteen studies used conventional non-simulation training methods as a comparator, such as lectures, surgical texts, or cadavers; three studies used alternative simulation-based training methods, such as simulation using mannequins and models; and one study used two comparator groups (one conventional and one simulation). In studies where iVR training was found to be more effective than comparator training methods, further analysis was conducted to ascertain the characteristics of successful iVR training interventions. This included an analysis of the duration of the intervention and the surgical specialty targeted by the training.

**Table 4:**
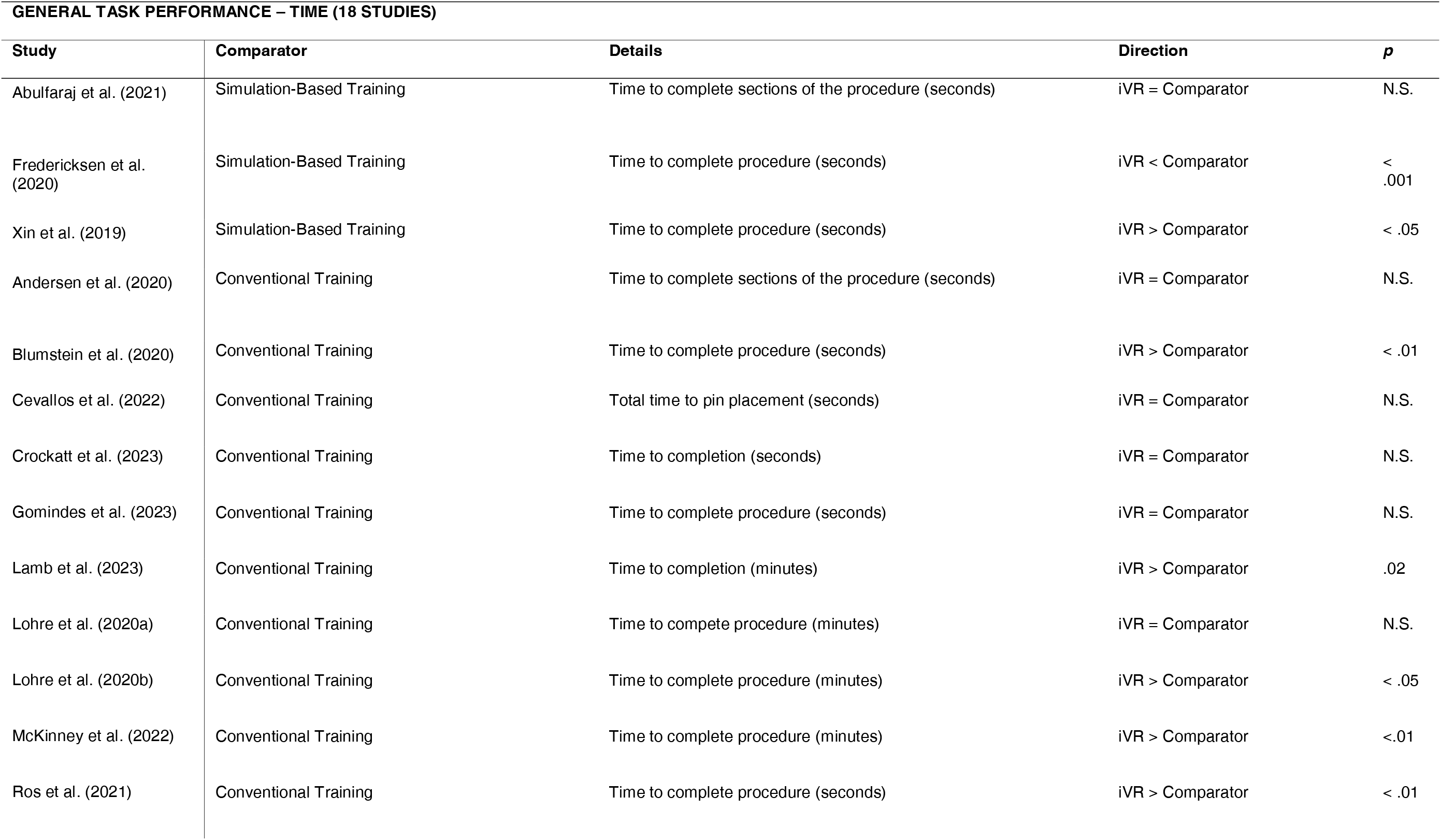
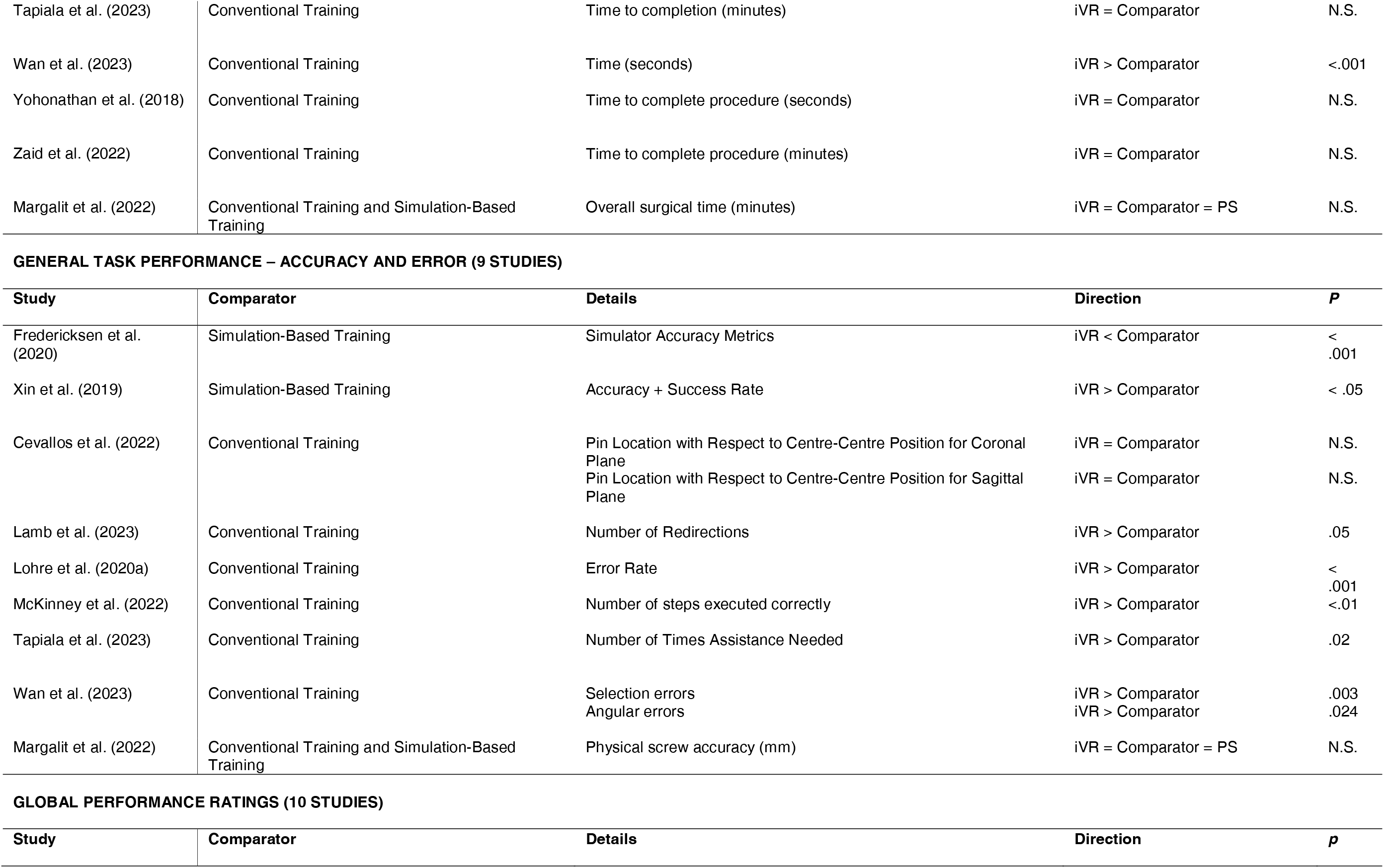
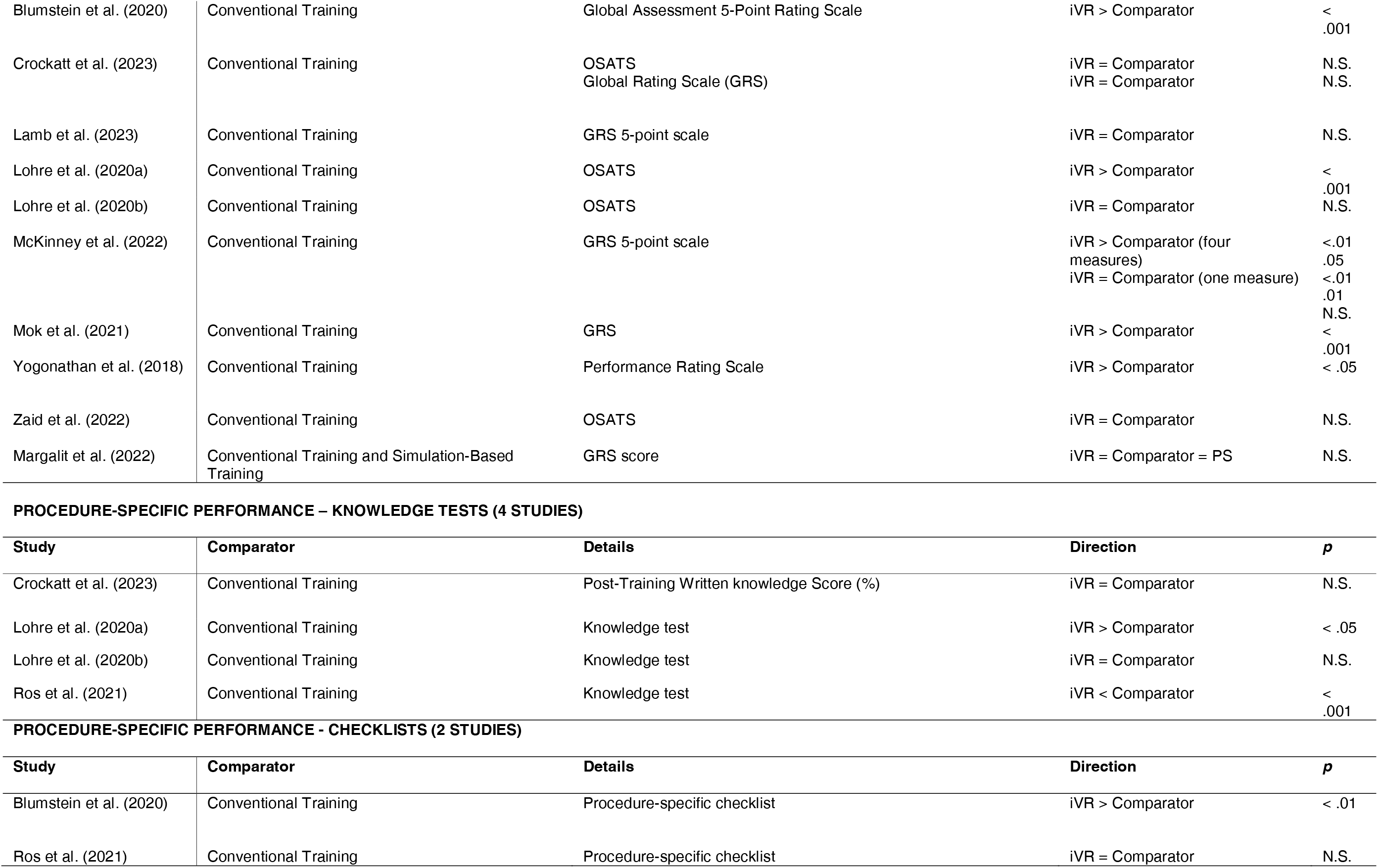
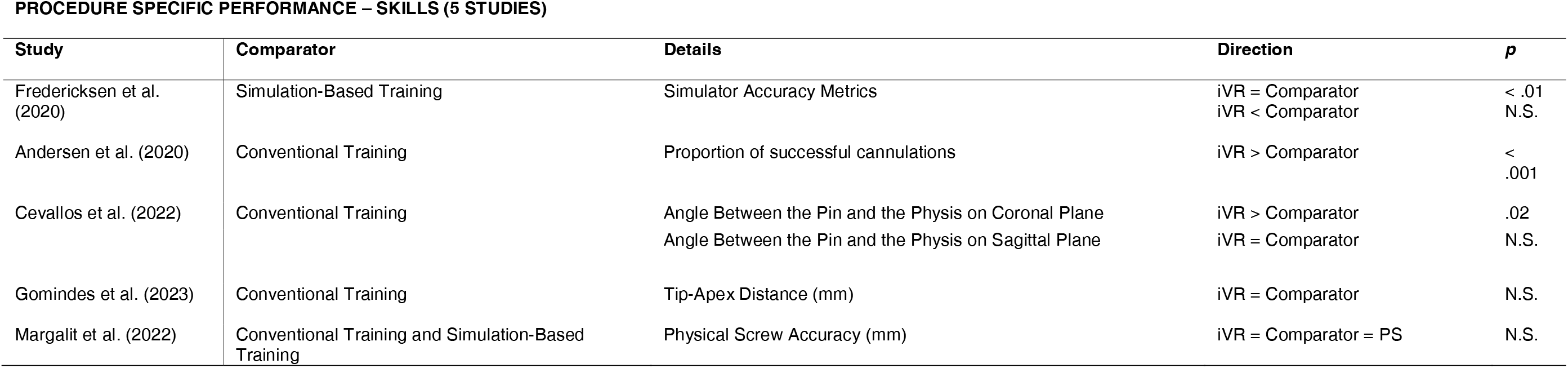
Studies grouped by outcome measure and comparator group.

GENERAL TASK PERFORMANCE – TIME (18 STUDIES)
| Study | Comparator | Details | Direction | p |
| --- | --- | --- | --- | --- |
| Abulfaraj et al. (2021) | Simulation-Based Training | Time to complete sections of the procedure (seconds) | iVR = Comparator | N.S. |
| Fredericksen et al. (2020) | Simulation-Based Training | Time to complete procedure (seconds) | iVR < Comparator | < .001 |
| Xin et al. (2019) | Simulation-Based Training | Time to complete procedure (seconds) | iVR > Comparator | < .05 |
| Andersen et al. (2020) | Conventional Training | Time to complete sections of the procedure (seconds) | iVR = Comparator | N.S. |
| Blumstein et al. (2020) | Conventional Training | Time to complete procedure (seconds) | iVR > Comparator | < .01 |
| Cevallos et al. (2022) | Conventional Training | Total time to pin placement (seconds) | iVR = Comparator | N.S. |
| Crockatt et al. (2023) | Conventional Training | Time to completion (seconds) | iVR = Comparator | N.S. |
| Gomindes et al. (2023) | Conventional Training | Time to complete procedure (seconds) | iVR = Comparator | N.S. |
| Lamb et al. (2023) | Conventional Training | Time to completion (minutes) | iVR > Comparator | .02 |
| Lohre et al. (2020a) | Conventional Training | Time to complete procedure (minutes) | iVR = Comparator | N.S. |
| Lohre et al. (2020b) | Conventional Training | Time to complete procedure (minutes) | iVR > Comparator | < .05 |
| McKinney et al. (2022) | Conventional Training | Time to complete procedure (minutes) | iVR > Comparator | <.01 |
| Ros et al. (2021) | Conventional Training | Time to complete procedure (seconds) | iVR > Comparator | < .01 |
| Tapiala et al. (2023) | Conventional Training | Time to completion (minutes) | iVR = Comparator | N.S. |
| Wan et al. (2023) | Conventional Training | Time (seconds) | iVR > Comparator | <.001 |
| Yohonathan et al. (2018) | Conventional Training | Time to complete procedure (seconds) | iVR = Comparator | N.S. |
| Zaid et al. (2022) | Conventional Training | Time to complete procedure (minutes) | iVR = Comparator | N.S. |
| Margalit et al. (2022) | Conventional Training and Simulation-Based Training | Overall surgical time (minutes) | iVR = Comparator = PS | N.S. |

GENERAL TASK PERFORMANCE – ACCURACY AND ERROR (9 STUDIES)
| Study | Comparator | Details | Direction | P |
| --- | --- | --- | --- | --- |
| Fredericksen et al. (2020) | Simulation-Based Training | Simulator Accuracy Metrics | iVR < Comparator | < .001 |
| Xin et al. (2019) | Simulation-Based Training | Accuracy + Success Rate | iVR > Comparator | < .05 |
| Cevallos et al. (2022) | Conventional Training | Pin Location with Respect to Centre-Centre Position for Coronal Plane | iVR = Comparator | N.S. |
|  |  | Pin Location with Respect to Centre-Centre Position for Sagittal Plane | iVR = Comparator | N.S. |
| Lamb et al. (2023) | Conventional Training | Number of Redirections | iVR > Comparator | .05 |
| Lohre et al. (2020a) | Conventional Training | Error Rate | iVR > Comparator | < .001 |
| McKinney et al. (2022) | Conventional Training | Number of steps executed correctly | iVR > Comparator | <.01 |
| Tapiala et al. (2023) | Conventional Training | Number of Times Assistance Needed | iVR > Comparator | .02 |
| Wan et al. (2023) | Conventional Training | Selection errors | iVR > Comparator | .003 |
|  |  | Angular errors | iVR > Comparator | .024 |
| Margalit et al. (2022) | Conventional Training and Simulation-Based Training | Physical screw accuracy (mm) | iVR = Comparator = PS | N.S. |

GLOBAL PERFORMANCE RATINGS (10 STUDIES)
| Study | Comparator | Details | Direction | p |
| --- | --- | --- | --- | --- |
| Blumstein et al. (2020) | Conventional Training | Global Assessment 5-Point Rating Scale | iVR > Comparator | < .001 |
| Crockatt et al. (2023) | Conventional Training | OSATS<br>Global Rating Scale (GRS) | iVR = Comparator<br>iVR = Comparator | N.S.<br>N.S. |
| Lamb et al. (2023) | Conventional Training | GRS 5-point scale | iVR = Comparator | N.S. |
| Lohre et al. (2020a) | Conventional Training | OSATS | iVR > Comparator | < .001 |
| Lohre et al. (2020b) | Conventional Training | OSATS | iVR = Comparator | N.S. |
| McKinney et al. (2022) | Conventional Training | GRS 5-point scale | iVR > Comparator (four measures)<br>iVR = Comparator (one measure) | <.01<br>.05<br><.01<br>.01<br>N.S. |
| Mok et al. (2021) | Conventional Training | GRS | iVR > Comparator | < .001 |
| Yogonathan et al. (2018) | Conventional Training | Performance Rating Scale | iVR > Comparator | < .05 |
| Zaid et al. (2022) | Conventional Training | OSATS | iVR = Comparator | N.S. |
| Margalit et al. (2022) | Conventional Training and Simulation-Based Training | GRS score | iVR = Comparator = PS | N.S. |

PROCEDURE-SPECIFIC PERFORMANCE – KNOWLEDGE TESTS (4 STUDIES)
| Study | Comparator | Details | Direction | p |
| --- | --- | --- | --- | --- |
| Crockatt et al. (2023) | Conventional Training | Post-Training Written knowledge Score (%) | iVR = Comparator | N.S. |
| Lohre et al. (2020a) | Conventional Training | Knowledge test | iVR > Comparator | < .05 |
| Lohre et al. (2020b) | Conventional Training | Knowledge test | iVR = Comparator | N.S. |
| Ros et al. (2021) | Conventional Training | Knowledge test | iVR < Comparator | < .001 |

PROCEDURE-SPECIFIC PERFORMANCE - CHECKLISTS (2 STUDIES)
| Study | Comparator | Details | Direction | p |
| --- | --- | --- | --- | --- |
| Blumstein et al. (2020) | Conventional Training | Procedure-specific checklist | iVR > Comparator | < .01 |
| Ros et al. (2021) | Conventional Training | Procedure-specific checklist | iVR = Comparator | N.S. |

**Table 4:** Studies grouped by outcome measure and comparator group.
| Study | Comparator | Details | Direction | p |
| --- | --- | --- | --- | --- |
| Fredericksen et al. (2020) | Simulation-Based Training | Simulator Accuracy Metrics | iVR = Comparator<br>iVR < Comparator | < .01<br>N.S. |
| Andersen et al. (2020) | Conventional Training | Proportion of successful cannulations | iVR > Comparator | < .001 |
| Cevallos et al. (2022) | Conventional Training | Angle Between the Pin and the Pysis on Coronal Plane<br>Angle Between the Pin and the Pysis on Sagittal Plane | iVR > Comparator<br>iVR = Comparator | .02<br>N.S. |
| Gomindes et al. (2023) | Conventional Training | Tip-Apex Distance (mm) | iVR = Comparator | N.S. |
| Margalit et al. (2022) | Conventional Training and Simulation-Based Training | Physical Screw Accuracy (mm) | iVR = Comparator = PS | N.S. |

### 3.4 Outcome Measures

#### 3.4.1 General Task Performance – Time (18 studies)

Time taken to complete either the whole taught procedure, reported by sixteen studies, or key sections of the procedure, reported by two studies, was the most common measure of general task performance. The findings for this outcome measure were mixed. Seven of the nineteen studies reported that iVR-trained participants were faster at completing the whole taught procedure than comparator participants. Blumstein et al. [43] found that the iVR group completed the Tibial Intramedullary Nailing procedure 147 seconds faster than participants who learnt the procedure by reading a surgical technique guide (p =.002). Lamb et al. [44] demonstrated that medical students trained using virtual reality completed the mock tibia IMN surgery 2.6 minutes faster on average than those trained using the traditional guide (p = .02). Lohre et al. [29] found that the iVR group was 7 seconds faster at completing the cadaveric glenoid exposure task compared to those who learnt the procedure using a journal article (p = .04). McKinney et al. [45] illustrated that participants who trained using immersive VR completed the unicompartmental knee arthroplasty 8.7 minutes faster than residents who trained using a technique guide (p < .01). Ros et al. [46] showed that the iVR group exhibited more efficient practical skills than the traditional lecture group, completing the lumbar procedure 74.5 seconds faster (p < .01). Wan et al. [47] found that medical students performed bimaxillary orthognathic surgery 279 seconds quicker if they were trained on the iVR surgical system instead of manuals and training videos (p < .001). Finally, Xin et al. [48] found that individuals in the iVR group were 126 seconds faster at placing pedicle screws in comparison to the standard video-trained group (p < .05).

However, a further eleven studies report that there was no difference in the time taken to complete either the whole procedure or key sections of the procedure. Abulfaraj et al. [49] reported that there was no significant difference in the time taken to correctly manage a simulated anaphylaxis and status epilepticus situation between participants who had trained using iVR and participants who had received training via a high-fidelity mannequin (p > .05). Anderson et al. [50] measured scanning time before peripheral venous catheter (PVC) insertion and time used for ultrasound (US) tip tracking and found no significant differences between the iVR group and the eLearning group (p >.05). Cevallos et al. [51] found no significant difference in the time taken for participants to pin a slipped capital femoral epiphysis between iVR training and a study guide (p = .26) Crockatt et al. [52] compared cadaver training and iVR training and found no significant difference in the time taken to complete augmented baseplate implantation during reverse total shoulder arthroplasty (p = .66). Furthermore, Gomindes et al. [53] discovered no significant difference in the time taken for surgical trainees to complete insertion of a short cephalomedullary TFNA nail into saw-bone femurs between traditional techniques and iVR training (p = .67). Lohre et al. [33] found that participants in the iVR group completed the reverse shoulder arthroplasty (RSA) procedure on a cadaver 492 seconds faster than those in the video-trained group (p = .13). Moreover, Margalit et al. [54] found no significant difference in surgical time between the reading and video group, the virtual reality group, or the physical simulation group for a slipped capital femoral epiphysis surgery simulation (p = .09). Tapiala et al. [55] trained medical students using either anatomy books or an iVR temporal bone model and found no significant difference in time to complete a mastoidectomy surgery between training methods (p = .50). In addition, Yoganathan et al. [56] found no significant difference between the iVR and the standard 2D video-trained group on time to completion (31 vs 30.5, p= 0.89). Zaid et al. [57] also found that there was no difference in the time taken to complete a knee arthroplasty on the SawBones simulator between their traditionally trained group, who learnt the procedure using a paper guide and an online demonstration video, and their iVR group (p = .9).

Only one included study reported that participants who were trained using iVR were slower than comparator participants. Frederiksen et al. [28] reported that the iVR group was slower at completing a laparoscopic salpingectomy than their comparator group (p < .001). Interestingly, this was the only study to compare iVR to standard, non-immersive VR.

#### 3.4.2 General Task Performance – Error Rate and Accuracy (9 studies)

Other measures of general task performance reported included error rates and accuracy. These measures favoured iVR over other training methods, with six out of nine studies reporting that iVR training reduced error rate and improved accuracy. Lamb et al. [44] measured the number of redirections required during the tibia IMN surgery and found that the conventional training group required significantly more redirections than the iVR training (iVR = 2.20 vs C = 2.50; p = .05). Lohre et al. [33] measured the error rate and found that the video-trained group made 50% more critical errors compared to the iVR group (65% vs 15%; P<.001). McKinney et al. [45] demonstrated that participants trained using VR executed significantly more steps correctly for a slipped capital femoral epiphysis surgery compared to conventional training (33 vs 27; p < .01). Additionally, Tapiala et al. [55] found that medical students trained on iVR required significantly less assistance during the mastoidectomy compared to those trained on anatomy books (10.70 vs 15.50; p = .02). Wan et al. [47] discovered that medical students trained on iVR made fewer errors during the orthognathic surgery compared to students trained on manuals and videos-both selection errors (1.00 vs 3.30; p = .003) and angular errors (7.90 vs 10.10; p = .024). Moreover, Xin et al. [48] found that the iVR group was more accurate than the control group (89.6% vs 60.4%; P < .05) and had a higher success rate (100% vs 79.2%, p< .05).

In contrast, Cevallos et al. [51] indicated that the pin location with respect to the Centre-centre position was not significantly different for iVR training and training with a surgical technique guide for either the coronal plane (2.55 vs 4.90; p =.46), or the sagittal plane (5.70 vs 4.95; p = .43). Frederiksen et al. [28], again the only study to compare iVR to standard VR, reported several accuracy metrics which were recorded by their simulator and, for all metrics, reported that accuracy was lower for iVR participants than for comparator participants (p < .001). Margalit et al. [54] also found no significant difference between iVR training and the comparator groups (reading/videos and physical simulation) for physical screw accuracy (iVR = 13.30mm, C = 24.4mm, PS = 16.3mm; p = .09) when participants completed an orthopaedic surgery.

#### 3.4.3 Global Performance Ratings (10 studies)

Evidence suggested that iVR training led to higher global performance ratings than other training methods, with five of the ten included studies reporting that iVR trained participants received higher ratings than comparators.

One common tool was the objective structured assessment of technical skills (OSATS), which was utilized by four studies, yet only one study found a significant effect. [34] Namely, Lohre et al [33] found that participants who received iVR training had significantly higher mean cumulative OSATS scores than the surgical video control group (15.9 vs 9.4; P<.001). In contrast, Lohre et al [29] found no significant differences between OSATS scores for the iVR group compared to the traditional learning group (11.8 vs 12.5; P= 0.70). Similarly, Zaid et al. [57] found no significant differences in OSATS scores for the iVR group compared to the standard training group (14.2 vs 15.7; P= 0.59).

Furthermore, eight studies included a unique global rating scale (GRS), four of which were statistically significant. Blumstein et al. [43] found that the iVR group showed superior results in all categories compared to the SG group (17.5 vs 7.5; P<.001), and this difference persisted after two weeks (19.9 vs 7.2; P<.001). McKinney et al. [45] reported that orthopaedic surgery residents significantly improved performance on a fixed bearing medial unicompartmental knee arthroplasty using iVR training compared to surgical guides for four of the five GRS categories (3.64 vs 2.36, p < .01; 3.73 vs 2.82, p = .05; 3.82 vs 2.54, p< .01; 3.45 vs 2.36, p = .01). Knowledge of specific procedures was the only category that was not significantly different between the training methods (3.64 vs 3.00, p = .12). In addition, Mok et al. [58] found that the iVR group performed better than the traditionally trained control group using two suture techniques: the Kessler method (24 vs 20, p<.001), and the Bunnell method (23 vs 20, p<.001) during a tendon repair procedure. Finally, Yoganathan et al. [56] included knot tying scores on a GRS designed for laparoscopic skills and found that the iVR group had a higher median compared to the video-trained group (5.0 vs 4.0; p = .04). However, Lamb et al. [44] found that iVR training did not significantly improve performance on tibia intramedullary nail (IMN) surgery compared to surgical guides (2.81 vs 2.68, p > .05). Margalit et al. [54] also found no significant difference between written material, physical simulation, and iVR training methods in terms of GRS for either reviewer for slipped capital femoral epiphysis surgery (p = .11, p = .06).

Additionally, one study utilised both OSATS and GRS ratings to measure performance, Crockatt et al. [52] found that participants trained on iVR did not have significantly different performance ratings compared to conventional training for augmented baseplate implantation during reverse total shoulder arthroplasty (OSATS: 91.20% vs 93.25%, p = .76; GRS: 4.71 vs 4.61, p = .70).

#### 3.4.4 Procedure-Specific Performance– Knowledge Tests (4 studies)

Of the four studies that included results from a knowledge test, only one reported that iVR trained participants scored higher on tests of procedure-specific knowledge than comparator participants. Lohre et al. [33] gathered verbal questioning scores relevant to reverse shoulder arthroplasty and found that those trained with iVR exhibited higher mean scores compared to those in the video-trained group (4.1 vs 2.2; P=.03). Conversely, Crockatt et al. [52] found no significant difference between junior residents post-training written knowledge score in those trained on iVR and those trained using cadavers for reverse total shoulder arthroplasty (58.30% vs 58.00%, p = .98). Lohre et al. [29], who conducted a knowledge survey relating to glenoid exposure as part of their study, found no significant difference between the iVR group and those who learned the procedure through traditional methods (13.5 vs 13.5, P=1). Additionally, Ros et al. [46] found that participants who were trained using iVR were significantly inferior to participants who received traditional lectures when required to complete an oral assessment consisting of eight theoretical questions relating to lumbar puncture (4.06 vs 4.97, p < .001).

#### 3.4.5 Procedure-Specific Performance – Procedure-Specific Checklists (2 studies)

Only two studies measured the number of steps on a procedure-specific checklist which participants completed. Blumstein et al. [43] found that participants within the iVR group successfully completed 38% more steps correctly of the procedure-specific checklist relevant to Tibial Intramedullary Nailing when compared to the Standard Guide group (63% vs 25%; P = 0.002). Ros et al. [46] found that there was no significant difference in the number of steps of a lumbar puncture procedure completed between participants in the iVR group and participants who had received traditional lectures (4.66 vs 4.77, p = .78).

#### 3.4.6 Procedure-Specific Performance – Procedure-Specific Skills (5 Studies)

Finally, five studies measured procedure-specific performance in terms of participants’ accuracy on procedure-specific skills. Andersen et al. [50] trained participants on ultrasound-guided PVC placement and found that iVR participants had a higher proportion of successful cannulations than comparator participants (0.73 vs 0.22, p < .001). However, Frederiksen et al. [28], again the study which compared iVR to standard VR, trained participants to complete a laparoscopic salpingectomy and reported that there was no difference in accuracy between participants in terms of the distance the tube was cut from the uterus (4.5 vs 4.6, p > .05). They also recorded a second measure of procedure-specific accuracy, diathermy damage, and reported that participants trained via standard VR caused less diathermy damage than iVR trained participants (1.3 vs 2.3, p < .01). Gomindes et al. [53] measured the tip-apex distance during insertion of a short cephalomedullary TFNA when investigating the difference between iVR training and Optech note training in residents and found no significant differences between the groups (9.00mm vs 15.00mm, p = .07). Additionally, Margalit et al. [54] found no significant difference in the physical screw accuracy for a slipped capital femoral epiphysis surgery between orthopaedic trainees who had iVR training, physical simulation training, and standard reading and video material training (13.30mm vs 7.70mm vs 9.4mm, p = .69). Finally, Cevallos et al. [51] compared iVR training to surgical technique guide training and measured the angle between the pin and the physis for a pinning of a slipped capital femoral epiphysis procedure. They found a significant improvement in the angle for iVR training when measuring on the coronal plane (2.55 vs 4.90, p = .02) however found no significant difference in training type for the sagittal plane (5.70 vs 4.95, p = .43).

### 3.5 Comparator Groups

#### 3.5.1 Conventional Training Methods (16 studies)

Of the sixteen studies which used a comparator group trained using conventional methods, such as lectures or surgical texts, twelve reported that iVR-trained participants performed better than the comparator group on at least one measure of surgical performance.[29,33,43–47,50,51,55,56,58] Only three studies, which included comparator groups trained using conventional methods, in these cases, surgical technique guides and cadavers, report no difference in all measures of surgical proficiency between the iVR and comparator groups.[52,53,57] One study utilised two comparator groups, one using conventional training and one using physical simulation training. This study reported only one measure where a significant difference was found between training groups-the number of radiographs taken. In this measure, iVR training was found to cause significantly fewer radiographs to be taken than either comparator group during a slipped capital femoral epiphysis model surgery.[54]

#### 3.5.2 Simulation-Based Training Methods (4 studies)

Findings for studies which included a comparator group that received simulation-based training were mixed. One study reported that participants trained using iVR performed better on outcome measures than participants who received simulation-based training using a structural spine model.[48] A second study reported no difference in performance between participants trained using iVR and participants trained using a high-fidelity mannequin simulation.[49] A third study, the only study to compare iVR to standard VR, reported that participants trained using iVR performed worse on both general task performance measures and procedure-specific measures than standard VR trained participants.[28] However, outcome measurements in this study were taken whilst participants used their assigned simulator type, which limits the comparisons that can be made as participants were not assessed in a standardised way. Finally, the study using both conventional and simulator training methods found only one significant difference between training method.[54]

### 3.6 Participants

#### 3.6.1 Medical Students (10 studies)

The majority of studies, eight out of ten, that used medical students in their participant group found that medical students who trained via iVR performed either the same as or significantly better than medical students in the comparator group on all recorded outcome measures. Andersen et al. [50] found that medical students who trained via iVR, compared to medical students who trained by watching a surgical video, completed significantly more successful cannulations during an ultrasound-guided PVC placement procedure on a phantom. There was no difference in the time taken for scanning prior to PVC insertion and for ultrasound tip tracking between the two groups. Blumstein et al. [43] reported that medical students trained via iVR to complete a tibial shaft fracture nailing procedure performed significantly better on all outcome measures when completing the procedure on a SawBones simulator than participants who learnt the procedure by reading a surgical technique guide. Cevallos et al. [51] reported medical students and surgical residents trained using iVR had a significantly more accurate angle between the pin and the physis on the coronal plane for a pinning of a slipped capital femoral epiphysis, however all other measures were not significant. Moreover, Lamb et al. [44] researched medical students either being trained using iVR or a standard surgical technique guide and found that students trained on iVR performed the tibia intramedullary nail surgery significantly quicker, and with fewer redirections required. However, the overall GRS score did not significantly differ between training methods. Another study, Mok et al. [58] found that the performance of medical students when completing a tendon repair procedure on a synthetic model was rated as significantly higher on the Global Rating Scale by expert surgeons for medical students who had trained via iVR compared to medical students who had learnt the procedure by watching a PowerPoint presentation. Tapiala et al. [55] found that medical students trained on iVR required less assistance than those trained on surgical guides for a mastoidectomy, although no other measure was significant. Wan et al. [47] reported that iVR training for medical students caused significantly greater performance on all measures of performance than technical manual training for orthognathic surgery. Finally, Xin et al. [48] reported that medical students who trained via iVR performed significantly better on all outcome measures when completing a pedicle screw placement on a cadaver than medical students who had trained via simulation on a structural spine model.

Two studies that used medical students as their participants did not find that iVR training clearly improved performance to a greater extent than other types of surgical training. Ros et al. [46] compared medical students trained to complete a lumbar puncture using iVR to medical students who learnt the lumbar puncture procedure by watching a lecture. Whilst medical students trained via iVR completed the lumbar puncture procedure on a mannequin significantly faster than medical students in the comparator group, medical students in the comparator group scored significantly higher on an oral assessment of their knowledge of the lumbar puncture procedure. There was no difference between the two groups of medical students in terms of the number of steps of the lumbar puncture procedure that they could remember correctly. Zaid et al. [57] included both medical students and residents in their participant groups. They found that there was no difference in performance on any outcome measure between participants who trained to complete a unicompartmental knee arthroplasty using iVR and participants who learnt the procedure by reading an illustrated surgical technique guide. For assessment, participants completed the procedure on a Sawbones simulator. The time taken to complete the procedure and rating on the OSATS scale were measured.

#### 3.6.2 Surgical Interns and Residents (11 Studies)

Six out of the eleven studies that used surgical trainees at the internship or resident level as their participant group found that participants who trained via iVR performed either the same as or significantly better than participants who trained via traditional surgical training methods on all outcome measures. Firstly, Yogonathan et al. [56] reported that the performance of foundation year doctors when completing a single-handed reef knot on a knot-tying jig was rated significantly higher on a performance rating scale by expert surgeons for foundation year doctors who had learnt how to complete the knot using iVR, compared to foundation year doctors who had learnt the knot by watching a 2D video. There was no difference between groups in terms of the time taken to complete the knot. Cevallos et al. [51] reported medical students and surgical residents trained using iVR had one significant result with other outcome measures not reaching significance, as stated above. Lohre et al. [33] found that senior orthopaedic surgery residents who learnt to complete a reverse shoulder arthroplasty using iVR, compared to those that had learnt the procedure by watching a surgical video, made significantly fewer errors when performing augmented glenoid implantation on a cadaver. They were additionally rated higher on the OSATS performance scale and scored higher on a knowledge test on reverse shoulder arthroplasty. There was no significant difference in the time taken to complete the procedure between groups. Additionally, Lohre et al. [29] compared orthopaedic surgical residents who learnt the glenoid exposure procedure using iVR to those that had learnt it by reading a technical journal article. Surgical residents trained via iVR were significantly faster than those in the comparator group when completing the glenoid exposure procedure on a cadaver. There was no difference between groups in terms of performance rating on the OSATS scale or performance on a test of knowledge about the procedure. Margalit et al. [54] measured performance on a slipped capital femoral epiphysis model surgery when orthopaedic trainees were trained using iVR, surgical guides, and physical simulation and found that trainees trained on iVR took significantly fewer radiographs during the mock surgery. No other measures of performance were found to be significant. Finally, McKinney et al. [45] compared iVR training to technique guide training for surgical residents for a fixed beading medial unicompartmental knee arthroplasty and found residents trained using iVR executed significantly more steps correctly and completed the procedure in a significantly shorter time period. Furthermore, they scored significantly higher on four out of the five GRS measures of surgical performance.

Conversely, four studies reported no difference between participant groups on all outcome measures. Abulfaraj et al.,[49] who compared medical interns trained to manage anaphylaxis and status epilepticus using iVR to medical interns trained using a high-fidelity model, found no difference in the time taken to complete critical actions when participants were required to manage a case of status epilepticus on a low-fidelity mannequin. Crockatt et al. [52] found no significant difference between residents who trained using iVR or cadavers for a reverse total shoulder arthroplasty on any measure of performance. Furthermore, Gomindes et al. [53] found no significant differences in performance for surgical trainees trained on either iVR or Optech notes for insertion of a short cephalomedullary TFNA. Zaid et al.,[57] who included medical students and residents in their participant groups, found no difference between groups on any outcome measure.

Finally, Frederiksen et al.,[28] the only study to compare iVR to standard VR, reported that first-year residents trained to complete a laparoscopic salpingectomy using iVR performed worse on both general task performance measures and procedure-specific measures than first-year residents trained via standard VR.

### 3.7 Characteristics of Effective iVR Interventions

#### 3.7.1 Surgical Specialty

The majority of effective iVR training interventions (eleven studies) focused on training participants to complete orthopaedic procedures, such as tendon repair or glenoid exposure. [29,33,43–45,48,51–54,58] Others focused on more basic surgical skills (two studies), such as tying surgical knots by using an ultrasound to place a peripheral venous cannula (PVC).[50,56] Two studies focused on other specialties; otolaryngology [55], and orthognathic surgery.[47]

#### 3.7.2 iVR Training Duration

For studies that reported the length of their iVR intervention in minutes, the duration of effective iVR training interventions ranged from 20 to 30 minutes.[43,48,50,53,56,58] Other effective interventions ranged from 1 to 2 hours.[45,51,52,55] Four of the effective iVR interventions allowed participants unlimited attempts at practicing with the iVR simulator, within a training session of an unspecified length.[29,33,44,54] One study allowed participants to complete ten sessions, each of which were 40 minutes long.[47]

### 3.8 Risk of Bias in Studies and Certainty of Evidence

The RoB (2.0) tool was used to assess the risk of bias in each of the studies for each outcome measure.[59] We assessed the methodological quality of included studies using the revised Cochrane ‘risk-of-bias’ tool for randomised trials, version 2.0 (RoB 2.0).[59] RoB 2.0 addresses five specific domains of potential bias: (1) bias arising from the randomisation process; (2) bias due to deviations from intended interventions; (3) bias due to missing outcome data; (4) bias in measurement of the outcome; and (5) bias in selection of the reported result. Two review authors independently applied the tool to each original included study and recorded supporting justifications for judgements, and the tool was applied by a researcher for each study included through the re-run of searches. Any discrepancies in judgements were resolved by discussion to reach a consensus. The RoB 2.0 tool provides an overall summary ‘risk-of-bias’ judgement (low; high; some concerns), for each individual outcome, and the overall RoB for each study was determined by the highest RoB level in any of the assessed domains, according to guidance by Higgins et al..[60] The majority of included outcome measures were judged, using the RoB 2.0 tool, to have “some concerns” relating to the overall risk of bias. This was primarily driven by bias in domain 5, selection of the reported result, and justified by researchers as no studies had a protocol with a pre-specified analysis plan. Only three outcome measures were classified as having a “high” risk of overall bias and, in all cases, this was due to biases in domain 1 (randomisation). “High risk” of bias judgements in this domain were justified by researchers as being due to a lack of clarity about the randomisation process, description of a randomisation process that was systematic, or failing to control for previous experience. Results for all other domains tended to be judged as “low risk”.

To assess for the certainty of evidence, we used the Mixed Methods Appraisal Tool (MMAT) version 2018.[61] The MMAT addresses five questions: (1) Is randomization appropriately performed? (2) Are the groups comparable at baseline? (3) Are there complete outcome data? (4) Are outcome assessors blinded to the intervention provided? (5) Did the participants adhere to the assigned intervention? The number of included studies was divided amongst the five researchers, and for each study, one researcher initially completed the MMAT with ‘yes’, ‘no’, or ‘can’t tell’ to each question, with justifications for their choice. Following this, a second researcher reviewed these responses, and in the case of disagreement, consensus was achieved through discussion. The curators of the MMAT discourage users to calculate an overall score from the ratings of each criterion, rather, it is ideally used to provide a more in-depth examination of each criterion.[61]

The MMAT additionally indicated that in most studies’ participant groups were judged to be comparable at baseline and participants adhered to their assigned intervention. However, there were issues with missing data across some studies, typically due to participants missing the assessment stage of the study, and that a minority of included studies did not state whether outcome assessors were blinded to participants’ interventions or whether randomisation was performed appropriately. The certainty of evidence assessment suggests findings should be interpreted with some caution.

## 4 DISCUSSION

Overall, data from objective performance metrics indicate that iVR training is more effective at improving surgical skill acquisition than conventional surgical training methods, such as surgical videos, journal articles, or cadavers, and as effective as other simulation-based training methods, such as simulation using a mannequin or model. These findings are consistent with those reported by previous systematic reviews, which have investigated the effectiveness of standard, non-immersive VR and found it to be as effective or more effective than other forms of surgical training.[16–20]

Interestingly, effective iVR training interventions were short in duration, with studies that specified the length of their iVR intervention describing training times that were between 20 minutes to 2 hours in length. This suggests that surgical trainees only need to access iVR training for a short period of time to benefit, indicating that surgical training programmes might need to purchase fewer iVR devices per trainee, thus increasing the cost-effectiveness of iVR as a training method. This would increase the viability of implementing iVR for many surgical training programmes.[7]

Additionally, analysis was performed to determine the effectiveness of iVR in improving different surgical outcome measures. For general task performance, seven studies found that participants in the iVR training group demonstrated significantly faster time to completion in comparison to traditional training methods,[29,43–48] whilst ten studies found no significant differences.[33,49–57] Moreover, iVR surgical training significantly improved accuracy and success rates [48,55] and reduced the number of critical errors made [33,44,45,47] when compared to non-iVR control groups. Hence, suggesting that iVR surgical training may be superior to non-VR alternatives for improving general task performance. These findings bear resemblance to a similar systematic review published by Mao et al. [25] which showed improved procedural times, task completion, and accuracy amongst iVR intervention groups.

Furthermore, the findings relating to global performance ratings are more congruent. All studies with results that were deemed statistically significant found that iVR surgical training was significantly more effective than non-VR training for measures of global performance.[33,43,45,56,58] Thus, it may be presumed that iVR is better than, or at least as good as, traditional learning methods, for surgical outcomes relating to global performance. This is comparable to a recent systematic review by Clarke et al. [62] of VR simulation in orthopaedic training, who found that in a large majority of the included studies, trainees using VR simulations outperformed those using traditional training methods on validated surgical skill checklists.

In terms of procedure-specific performance, four studies found significant effects in favour of iVR,[33,43,50,51] compared to six studies that found no significant difference.[28,29,46,52–54] On the other hand, Ros et al. [46] found that participants in the traditional lecture group showed statistically significant improved performance during the oral assessment in comparison to the iVR group. This suggests that iVR may be effective for improving procedural knowledge, but less so for declarative knowledge, a phenomenon also noted by Mao et al. [25]. This may be due to iVR training having a greater focus on the psychomotor skills which are required to perform the procedures, rather than the technical theory behind the procedure.

Interestingly, iVR surgical training may be less efficacious than its predecessor, as Frederikson et al. [28] found that a control group trained via non-immersive VR significantly outperformed the iVR group in all metrics, and this was presumed to be due to the increased cognitive load imposed by more immersive technologies. Nevertheless, this finding does not negate the potential usefulness of iVR; rather, Frederikson et al. [28] recommend introducing iVR in surgical skills training after initial training using CVR, to bridge the gap between simulation and real-life operating room scenarios.

### 4.1 Limitations

#### 4.1.1 Limitations of the Evidence

There is high heterogeneity between studies, with a broad range of surgical disciplines, comparators, and outcomes, making it difficult to draw definite conclusions. Furthermore, procedure-specific outcomes, by their nature, are difficult to generalise to other surgical disciplines. This limits our ability to determine if iVR training is more effective for certain surgical procedures, which could explain the diversity in results. Additionally, samples were small and iVR intervention duration in the included studies was short, with training lasting less than a day. However, core surgical training amongst medical students can last up to two years, reducing the ecological validity of the findings. Therefore, the effects of iVR training throughout the entire residency programme are still to be elucidated. Contrastingly, as most studies utilised cadavers or simulators to test the surgical skill trained, they may not represent the environment of the surgery on an actual patient, limiting the generalisation of iVR training to real patients. However, cadavers have proven skill transfer validity suggesting this method of testing skill acquisition may be adequate.

#### 4.1.2 Limitations of Review Processes

As many of the included studies reported no measure of variation, it was impossible to calculate effect size estimates. Due to this, vote counting based on the direction of effect was used for synthesis, with further elaboration with p-values, both of which provided no information about the magnitude of the effect of iVR training on surgical performance. This means that, although we know that iVR is as effective or more effective for training surgical trainees than more conventional training methods, we do not know the size of the effect that iVR training has on surgical performance.

#### 4.1.3 Implications

Considering that iVR has emerged as a potentially effective method of surgical training provision, this has implications for practice and policy, as it highlights the viability of iVR simulators within surgical residency curricula to supplement learning, which could reduce reliance on traditional apprenticeship models and cadavers. Due to their lower cost, commercially available iVR devices mean that the widespread integration of iVR into surgical training programmes is more viable.[7] Viability is further increased by the finding that surgical trainees only need access to iVR training for short periods of time before improvements in their surgical skills are found.

In addition, iVR could be utilized as an objective assessment tool for procedural-based specialities, with the possibility of adapting surgical board certification examinations.[25] Furthermore, iVR could be used to aid surgical skill development in areas of limited access, so that individuals can gain the necessary expertise whilst avoiding the associated risks. Nevertheless, before iVR is integrated into real-world surgical training, we recommend further investigation through large-scale studies with long follow-up periods, possibly focusing on specific surgical domains, to address the foregoing critiques and provide a fully enlightened consensus on the efficacy of these systems.

## 5 RECOMMENDATIONS FOR FUTURE RESEARCH

We end this review by offering 10 recommendations for future research on iVR in surgical training, to provide a roadmap for conducting more comprehensive and rigorous research in this area. These recommendations touch on various aspects of iVR research in surgical training, including the need for standardization of definitions and reporting, the exploration of different outcome measures, the investigation of long-term impacts and cost-effectiveness, and the examination of individual differences among trainees. By following these recommendations, we believe that a more nuanced understanding of the potential of iVR as a surgical training tool can emerge. This could ultimately contribute to the development of more effective and efficient surgical training programmes, leading to improved surgical performance and (ultimately, having positive consequences on) patient outcomes.

1. Standardization of iVR Definition: Our review underscores the need for a clear and standardized definition of iVR in the context of surgical training research. It was observed that many studies incorrectly labelled their training intervention as “immersive” without using a head-mounted display (HMD) or an interactive interface, which are key components of true iVR. Therefore, it is recommended that future research adopts a standadrised definition of iVR used in this review to ensure consistency and accuracy in the field. This would facilitate more precise comparisons across studies and contribute to a more coherent and robust body of literature on iVR in surgical training.
2. Detailed Reporting of iVR Training Interventions: Going beyond nomenclature, future research should aim for detailed and standardized reporting of iVR training interventions. This includes specifics about the iVR equipment used, the design of the iVR simulations, the duration and frequency of training sessions, and the specific surgical skills targeted. This level of detail would allow for a more nuanced understanding of the factors that contribute to the effectiveness of iVR training and facilitate more accurate comparisons across studies. Wider use of existing reporting guidelines for simulation-based medical research may support this goal [63,64].
3. Conducting Larger Scale, Longitudinal Studies: Procedure-specific outcomes are difficult to generalize to other surgical disciplines, and the short duration of iVR interventions in the included studies, coupled with small sample sizes, further complicates the interpretation of results. Future research making use of large-scale, longitudinal studies that span the entire duration of surgical residency programmes would provide a more comprehensive understanding of the effectiveness of iVR training. Within these studies, future research could include more comparative studies to better understand the relative effectiveness of iVR compared to other advanced simulation-based training methods. This would help to position iVR within the broader landscape of surgical training tools.
4. In-depth Analysis of Experience Surveys: While the review considers experience surveys as a secondary outcome measure, future research could delve deeper into the subjective experiences of surgical trainees. This could involve qualitative studies or more detailed surveys to gain insights into trainees’ perceptions of iVR training, its usability, and its impact on their confidence and motivation.
5. Investigation of Learning Rate and Mechanisms: Few of the reported studies attempted to characterise learning over time, and little attention was given to the cognitive processes supporting that learning. Understanding how quickly and effectively surgeons can acquire new skills using iVR and could provide valuable information on the optimal duration and frequency of iVR training sessions. Longitudinal measurement of performance changes during iVR training could help to support this. More generally, to maximise the benefits of iVR training, more basic research into the mechanisms of sensorimotor skill learning and their interactions with higher-level declarative knowledge will be required.
6. Integration with Other Training Methods: While this review compares iVR with other training methods, future research could explore how iVR can be integrated with other training methods to create a comprehensive hybrid surgical training programme. This could involve a combination of lectures, hands-on practice with cadavers or surgical models, and iVR simulations. This would help to understand the synergistic effects of different training methods and provide a more holistic approach to surgical education.
7. Impact of iVR on Teamwork and Communication Skills: Most iVR training experiences involve a lone performer, a drastically different situation from the one found in the operating room. Future research could explore the impact of iVR training on non-technical skills such as teamwork and communication, which are crucial in surgical settings. This could involve designing iVR simulations that require collaboration and communication among trainees.
8. Exploration of Individual Differences: Future research could also explore how individual differences among surgical trainees, such as prior experience with virtual reality and general sensorimotor aptitude, influence the effectiveness of iVR training. This could provide insights into how to tailor iVR training to individual trainees to maximize its effectiveness. There are also known accessibility issues for specific groups and this topic requires deeper consideration if these technologies are to be scaled across the surgical workforce.
9. Cost-Benefit Analysis: While the potential cost-effectiveness of iVR as a method of surgical training is often mentioned, more comprehensive cost-benefit analyses are required for organisations to be able to calculate the potential return on investment. Some parts of this are relatively straightforward to calculate (cost of equipment, training time) but there are other, more intangible benefits (e.g., accessibility, stress reduction, confidence) that need consideration too.
10. Effect of iVR on Patient Outcomes: Lastly, future research will need to investigate the impact of iVR training on real-world patient outcomes. Improved patient outcomes are ultimately the end goal of these training initiatives. Capturing the impact of training is, of course, a difficult challenge, but as this field grows, there needs to be a concerted effort to develop approaches that can start to quantify the impact of training in this way.

## 6 CONCLUSIONS

Our systematic review comprehensively examines the role of immersive virtual reality (iVR) in skill acquisition. The findings suggest that iVR training can be an effective tool for improving surgical performance, with potential benefits over conventional training methods and comparable effectiveness to other simulation-based methods. However, the review also highlights several areas for further investigation, including the long-term impact of iVR training, the influence of individual differences among trainees, and the integration of iVR with other training methods. The review underscores the need for more rigorous and standardised research in this field, with a particular emphasis on including measures of variation, conducting large-scale longitudinal studies, and providing detailed reporting of iVR interventions. By addressing these areas, future research can contribute to a more robust and nuanced understanding of the potential of iVR in surgical education.

## Data Availability

All data produced in the present work are contained in the manuscript

## ACKNOWLEDGEMENTS

This research is funded by the National Institute for Health and Care Research (NIHR) Leeds Biomedical Research Centre (BRC) (NIHR203331). The views expressed are those of the authors and not necessarily those of the NIHR or the Department of Health and Social Care. Author FM is further supported by the European Union’s Horizon Research and Innovation programme under grant agreement no. 101070155 and the UKRI through the Horizon Europe Guarantee (#10039307). The authors would like to thank the University of Leeds School of Psychology MSc students who helped facilitate the data undertaking of the systematic review.

